# The Impact of Face-Masks on Total Mortality Heterogenous Effects by Gender and Age

**DOI:** 10.1101/2021.06.08.21258545

**Authors:** Giacomo De Giorgi, Maria Maddalena Speziali, Felix Michalik

**Affiliations:** IEE/GSEM University of Geneva BREAD, CEPR, and IPA; IEE/GSEM University of Geneva Universitá Magna Graecia of Catanzaro; BGSE

## Abstract

Governments around the world have been implementing several non-pharmaceutical interventions (NPIs) to fight Covid-19 spread and its associated mortality. We estimate the causal impact of mandatory face-mask wearing policy in public places on (total) mortality in Switzerland. We exploit the staggered introduction of the policy across Swiss cantons using a Difference-in-Difference and an event study approach. We find that the extension of compulsory mask wearing to public places has an heterogeneous impact on mortality, with small positive effects on male mortality entirely driven by older age-cohorts (90+). Finally, we show that adding contact tracing and stricter distancing to compulsory face-mask policy does not lead to better results in terms of mortality.

## 1 Introduction

By the end of 2019 China, then Europe, and the rest of the world have lived in a constant crisis mode due to COVID-19. On January 30, 2020 the World Health Organization (WHO) declared the outbreak as a “Public Health Emergency of International Concern”. To contain the spread of the virus, governments have resorted to several Non-Pharmaceutical Interventions (NPIs) including distancing rules, tracing, mandatory face-masks, and partial or total lock-downs. Vaccination policy in European countries is still far from achieving “herd immunity”, as of May 18, 2021 only about 27% of European population has received at least the first dose of vaccine. Thus, public health policies on NPIs remain the prime tools to prevent the spread of contagion and associated deaths.

However, after more then a year since the start of the pandemic, the debate on which combination of NPIs better contains Covid-19 spread is still burgeoning (Mei, 2020). The existing literature mostly agrees on the positive effects of distancing and lock-down policies (Courtemanche et al., 2020; Pei et al., 2020; Andersen, 2020; Abouk and Heydari, 2020; Nadig and Krishna, 2020; Thunström et al., 2020; Conyon et al., 2020), while lower consensus exists on the effectiveness of face-masks for reducing mortality.

In this paper we contribute to the ongoing debate on the effectiveness of mandatory face-mask policy. In particular, we estimate the effect of compulsory wearing of face-mask in Switzerland over the first wave of pandemic in a quasi-experimental setting looking at overall mortality as our main outcome (see also Salathé et al., 2020, Nivette et al., 2020). We find a small and heterogeneous effect across gender and age-class, with a 7.5% reduction in total mortality for males only (see Haischer et al., 2020 for an observational study in Wisconsin, US).

The existing literature mainly focuses on covid-related outcomes to evaluate NPIs success. In particular, the growth rate of covid deaths and confirmed cases (Chernozhukov et al., 2020; Courtemanche et al., 2020; Mitze et al., 2020; Lyu and Wehby, 2020; Conyon et al., 2020; Brauner et al., 2021; Bendavid et al., 2020) and covid transmission rate, *R*_*t*_ (Pei et al., 2020; Flaxman et al., 2020; Avery et al., 2020). These outcomes are of great interest, they may however suffer from mis-measurement as in any epidemic and in particular in a new one for the first wave (Alicandro et al., 2020). For example, it can be hard to distinguish between people dead *because of* Covid-19 and those who died *with* Covid-19 or simply a number of deaths from Covid would go undetected, or mortality may be affected from untreated conditions. It is therefore of great interest to look into total mortality as the measure will capture also deaths from untreated conditions, further to being less prone to measurement concerns.

Two key aspects on Covid containment policies in Switzerland are instrumental to our analysis. First, over the initial months of the pandemic, the different cantons had substantial autonomy over the Swiss Federal Council. As a result, public health policies differed across cantons. Thus, throughout the paper we distinguish between Federal and Cantonal policies. Where, the former were imposed by the Swiss Federation and applied to all cantons; while the latter were decided by a specific cantonal authority and accordingly enforced within the canton only. Secondly, on July 6, 2020, the Federation mandated face-mask wearing on public transportation. Then, between July and October, 9 cantons extended face-mask requirements to all public (indoor) places, e.g. supermarkets, stores, and restaurants. Our object of interest is the effect of extending face-mask requirements to these extra locations. Then on October 18, the Federation applied the stricter mask rules to the whole country.

An increasing number of studies in different fields is now focused on face-masks ability to prevent Covid-19 infections (Howard et al., 2020; Karaivanov et al., 2020; Mitze et al., 2020; Lyu and Wehby, 2020; Haischer et al., 2020) and how filtering capacity differs across types of masks (see Long et al., 2020; Howard et al., 2020; Leung et al., 2020; Bae et al., 2020; Li et al., 2020). How do cultural factors and governments’ approach to combat the pandemic affect the portion of population wearing mask (Haelle, 2020 and Feng et al., 2020). Does the positive association between mask wearing and the reduction of viral spread arise through protection of uninfected wearers (protective effect), via reduced transmission from infected mask wearers (source control), or both (Bundgaard et al., 2020; Howard et al., 2020)? How do we measure policy compliance (Chernozhukov et al., 2020; Haischer et al., 2020)? Estimating the effectiveness of mandatory face-mask policy (and NPIs in general) is hard for at least three reasons: (i) unreliable data; (ii) lack of credible sources of variation in the adoption of the different measures; and (iii) large heterogeneity in the results (Stock et al., 2020).

We overcome such issues with a novel approach on both data and methods. First, our analysis is based on a set of reliable administrative data. We construct a panel containing data on the weekly number of deaths in each Swiss canton between 2000-2020. We also collect data on total population between 1999-2019. With this information we calculate the *‘weighted total deaths’*. Using this outcome allows us to overcome the measurement issues that are likely to affect reported data on covid deaths or typical of survey methods. Second, the cantons’ autonomy in policy making gives us a reliable source of variation in the intervention. Thus, we estimate the impact of additional face-mask requirements with a quasi-experimental approach. Namely, we employ a Difference-in-Difference and an Event-study approach. Our empirical strategy is new for two reasons. First, we set the post policy period from the date mask wearing policy is introduced in the first canton (July 7). This allows us to account for likely anticipation effects due to people behaviors and information flows. Second, with long data series on deaths we can control for canton and time fixed effects (week of the year for example) and allow for a very flexible pre-trend in the outcome variables. Further, we can also look at how the effect of the policies evolves over a number of weeks up to 40 weeks since implementation (dynamic effects). Our estimates show a small and insignificant effect overall across age and gender, both for the Diff-in-Diff and event study approaches, with a marginally significant and small decrease in the mean of deaths for male (7.5% - significant at the 10% level) in the main specification, but no effect in the event study and the staggered Diff-in-Diff (where we attribute the implementation of the policy at exactly the date the specific canton enacted the stricter mask-wearing requirements). For female mortality we find a negligible and imprecisely estimated increase (2.2% -*p* − *value >* .5 in the main Diff-in-Diff). Finally, we investigate two likely sources of heterogeneity in the results. First, we conduct an analysis based on demographic characteristics. It is well-known that the infection fatality rate (IFR) varies dramatically along the age and gender dimensions. For people in their mid-seventies or older the IFR is attested to be around around 11.6% (Mallapaty, 2020). Gender is also associated with Covid-19 mortality. Men are almost twice as likely as women to die from the coronavirus (Pastor-Barriuso et al., 2020). To analyse the demographic heterogeneity, we provide detailed evidence on 4 age groups: young (0-29), middle (30-59), old (60-90) and very old (90+). We estimate the effects for each group separately, for males and females, and in a pooled-regression. The analysis confirms a positive effect (lower mortality) on males, driven by the oldest age-class, with a decrease of 17% − *pvalue <* .1 in the 90+ deaths. While, on females we find a null effect on the outcome for all age classes. The second likely source of heterogeneity in the results comes from the mix of mandatory face-masks with other containment measures. Indeed, timing and combination of NPIs adoption may affect their performance (Markel et al., 2007; Willi et al., 2020). Flaxman et al., 2020 analyze several policies (lockdown, public event limitation, school closures, self isolation and distancing encouragements) and find lock-down to be the most effective. In contrast, Brauner et al., 2021 argue that the additional effect of stay-at-home orders was comparatively small with respect to shuttering educational institutions, limiting gatherings to 10 people or less and closing face-to-face businesses. Bendavid et al., 2020 provide evidences on the small effect of stricter measures (stay-at-home order and business closure) when less restrictive NPIs have been already enforced (e.g. discouraging of international and domestic travel, and a ban on large gatherings). Within our policy-mix analysis we examine the effect of additional restrictions with respect to the simple Federal requirement of face-masks on public transport. Namely, we estimate the effects of combining stricter masks requirement with two further NPIs: (i) distancing and (ii) tracing policies. The policy-mix analysis confirms the overall small effect of the policy and its heterogeneity across genders. Indeed, we find stricter face-mask requirement to be more effective than Federal policy only on male mortality in the DiD design. While, female population has not additional benefits from different combinations of policies. Further, the analysis suggests that face-mask policy ‘s achievements cannot be disentangle from distancing and tracing effects.

The reminder of the paper is organized as follow. In Section 2 we provide a chronology of Federal and cantonal policies. In Section 3 we describe the data and the outcomes of interest. In Section 4 we report our key identification strategy and the main results. Section 5 shows that the results are robust to a series of checks. Finally, in 6 we explore two likely source of heterogeneity in our results. Conclusions are drawn in 7.

## 2 Policy Timeline

Over the first wave of the pandemic, Covid-19 containment policies in Switzerland were enacted on two levels. Federal policies, imposed by the Federal Council, were valid in the whole country. While Cantonal policies, decided by Cantonal Authorities, applied to the specific canton only.

### Federal Policies

On March 16 2020, the Federal Council declared Switzerland to be in an ‘Extraordinary Situation’, imposing a partial lock-down.^1^ After a month the Federal Government announced an easing of the restrictions in small steps between April and July. Hence, between April 27 and the end of June, most of the businesses, bars and restaurants reopened. People were always required to leave their contact information in written form (e.g. name and telephone number). Primary and lower secondary schools - as well as higher levels of education in groups of 5 - were resumed. Starting by June 6 clubs, cinemas and all other leisure activities were restored. Also events with a number of participants between 300 and 1000 were allowed again. In the whole country, as of July 6 wearing a face masks became compulsory in public transportation and quarantine rules were introduced for travellers coming from regions with high risk of infection.^2^ Finally, on October 18, 2020 the Federation introduced a mandate of face-mask wearing in all public indoor spaces (e.g. supermarkets, train stations, libraries, shops).

By the beginning of July some of the cantons started to impose their own policies on NPIs, including distancing, tracing and stricter requirements on face-mask wearing. Some of the cantons did not take additional measures beyond those imposed by the Federation in that entire period (Appenzell Ausserrhoden, Appenzell Innerrhoden, Glarus, Nidwalden, Obwalden, Switz, Uri).

### Aargau

From July 3 onward, the canton has required clubs, bars and restaurants to gather contact details of their guests. By July 9, the canton has limited the maximum number of guests in restaurants and events to 100.

### Basel-Landschaft and Basel-Stadt

From July 6 onward the cantons have required clubs, bars and restaurants to gather contact details of their guests. By July 9, the cantons have limited the maximum number of guests in restaurants and events to 100. In addition, on August 24 **Basel-Stadt** made face-mask mandatory in shops and for restaurant employees.

### Bern

From July 17 onward the canton has required clubs, bars and restaurants to gather contact details of their guests. By October 12, the canton has limited the maximum number of guests in restaurants and events to 100. In addition, customers must consume food and drinks while sitting at their tables. On October 12 Bern made face-mask mandatory in all public indoor spaces.

### Fribourg

From July 17 onward the canton has required clubs, bars and restaurants to gather contact details of their guests. In addition, on August 24, Fribourg made face-mask mandatory in shops and restaurants, for both guests and employees.

### Geneva

From July 24 onward the canton has required clubs, bars and restaurants to gather contact details of their guests. In addition, on July 24 Geneva made face-mask mandatory in restaurants, for employees only. The canton extended the requirement to shops on July 28, and to bars and restaurants guests on July 31. Starting from July 31 clubs have closed. From August 18 onward the canton has required bars and restaurants to gather contact details of their guests. Further, private events were allowed up to 100 attendees, public events of up to 1000 participants had to be divided into sections with up to 100 people each. Finally, on October 14, Geneva made face-mask mandatory in all public places.

### Grischun

On October 17, the canton made face-mask mandatory in all public indoor spaces.

### Jura

On July 7, the canton made face-mask mandatory in shops. From August 25 onward the canton has required clubs, bars and restaurants to gather contact details of their guests. The canton extended the face-mask requirement to restaurants staff and customers and spectators of indoor events on October 9. By the same date, the canton has limited the maximum number of people in private events to 100. Since October 14 these measures have also applied to private events and tracing of guests must be done electronically.

### Luzern

From July 4 onward the canton has required clubs, bars and restaurants to gather contact details of their guests. By July 17, the canton has limited the maximum number of guests in restaurants and events to 100. Public events of up to 1000 participants had to be divided into sections with up to 100 people each and information of attendees had to be gathered. On October 17 Luzern made face-mask mandatory in all public indoor spaces.

### Neuchâtel

On August 21, the canton made face-mask mandatory in shops (except if less then 10 people were in the shop). By the same date, the canton has limited the maximum number of guests in restaurants and events to 100.

### Schaffhausen

From July 10 onward the canton has required clubs, bars and restaurants to gather contact details of their guests. On October 16, Schaffhausen made face-mask mandatory in shops and supermarkets.

### Solothurn

From July 3 onward the canton has required clubs, bars and restaurants to gather contact details of their guests. By July 9 the canton has limited the maximum number of guests in restaurants and events to 100. On September 3 Solothurn made face-mask mandatory in all shops.

### St. Gallen

From September 25 onward the canton has required clubs, bars and restaurants to gather contact details of their guests.

### Thurgau

From August 14 onward the canton has required clubs, bars and restaurants to gather contact details of their guests.

### Ticino

By July 9 the canton has limited the maximum number of people in public spaces to 100. On October 9 all clubs have closed. On October 10, Ticino made facemask mandatory in shops. From the same date onward the canton has required clubs, bars and restaurants to gather contact details of their guests. At a time, restaurants’ guests have been allowed to drink and eat while seated.

### Valais

By July 16 the canton has limited the maximum number of guests in restaurants and events to 100 (after 8 PM). From the same date onward the canton has required clubs, bars and restaurants to gather contact details of their guests. On August 31, Valais made face-mask mandatory in all shops.

### Vaud

On July 8 the canton made face-mask mandatory in all shops providing that there were more than 10 people. From the same date onward the canton has required clubs, bars and restaurants to gather contact details of their guests. On September 17 the canton extended the face-mask requirement to restaurants’ staff and customers. At once Vaud introduced a number of additional measures: all clubs had to close, the maximum number of guests in restaurants and events was limited to 100, while for events with more than 50 people masks were mandatory (organizers must also keep a list of participants). Finally, from the same date onward the canton has required clubs, bars and restaurants to gather contact details of their guests.

### Zug

From July 13 onward the canton has required clubs, bars and restaurants to gather contact details of their guests. By July 13 the canton has limited the maximum number of guests in restaurants and events to 30. On August 22 Zug introduced a number of additional measures: events with more than 100 attendees were only allowed if distancing could be maintained or masks worn. At the same time measures for bars and clubs were relaxed (100 people were allowed indoors). On October 10, Zug made a face-mask mandatory in shops, supermarkets and restaurants.

### Zürich

From July 3 onward the canton has required clubs and bars to gather contact details of their guests. The requirement was extended to restaurants from August 27. By the same date, the canton made face-mask mandatory in all shops and supermarkets and also limited the maximum number of guests in restaurants and events to 100 (rising to 300 provided that an outdoor area was allowed). For private events (e.g., weddings), concerts or church events with more than 100 participants the mask mandate applied. As of September 24 restaurants, bars, and clubs, in which consumption was not necessarily done while seated, were allowed to host up to 300 guests again, as long as face masks were kept. Moreover, prostitutes were required to collect and verify the contact details of their customers. On October 15, Zürich made face-mask mandatory in all restaurants, bars and clubs.

**Figure 1:**
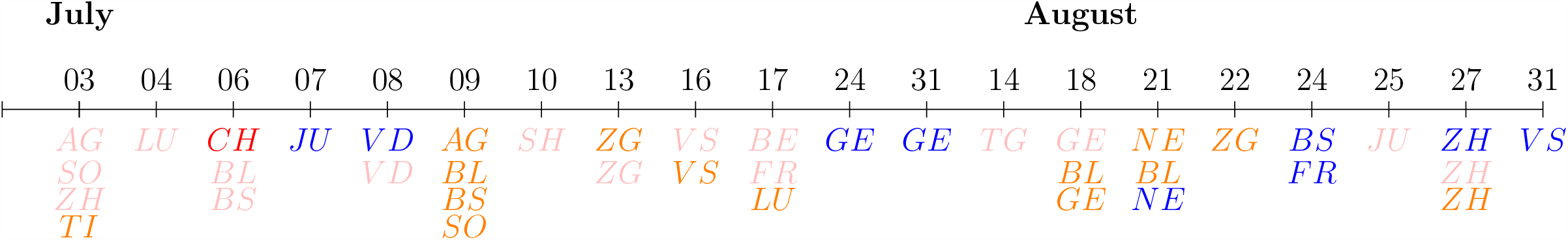
Policy Timeline (July-August)

**Figure 2:**
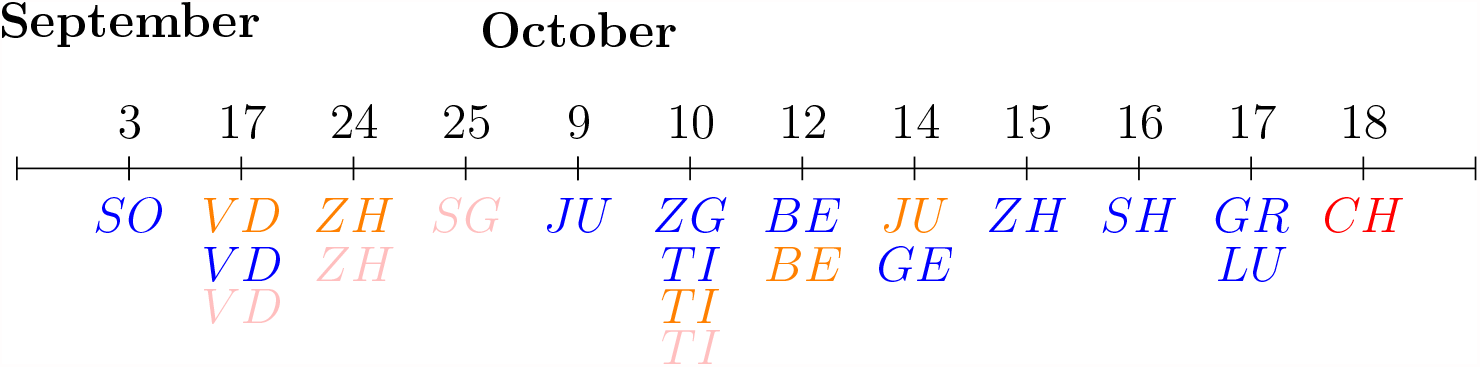
Policy Timeline (September-October) Note: Policies are grouped in 4 categories. In red mask requirements imposed by the Federation, involving all the cantons, are reported. In pink tracing policies are reported. In Orange social distancing policies are reported.(e.g. by limiting the number of people to 1000 in public events, 100 or 10 in private ones). Finally in blue Mask Policy are reported.

## 3 Data and Outcome

Our analysis is based on a longitudinal dataset at the canton-week-year level. The dataset comprises observations for the 26 Swiss cantons in the first 40 weeks of each year between 2000 and 2020. Even if the post-policy period ends in week 42 of year 2020 (October 18), we cut our analysis two weeks earlier. This refinement arises because five cantons applied the stricter requirements in those two weeks, i.e. October 10^*th*^ for Zug and Ticino, October 12^*th*^ for Bern, and October 17^*th*^ for Grischun and Luzern.

The final database combines several administrative sources. To construct the policy timeline we use information from newspaper articles and cantonal official internet web-pages. Data on total deaths and population comes from the Federal Statistical Office website, while information on Covid deaths and cases is gathered from Zurich Statistical Office.^3^ For both deaths and population we have information on gender (Male and Female) and age in 5-years intervals. We then group ages in 4 cohorts (0-29; 30-59; 60-90; 90+) according to WHO classes. Using this information we compute the main outcome of interest as follow:

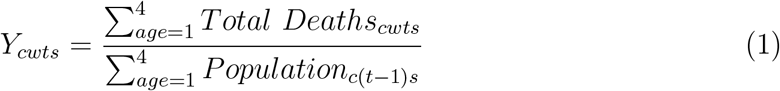

Where, *Y*_*cwts*_ represents the share of deaths in canton *c*, in week *w* and year *t* for gender *s*. The aggregate number of male (female) deaths is weighted with canton’s male (female) population in the previous year (*t* − 1). This adjustment allows us to take into account (i) the different population sizes of cantons (ii) cantonal demographic characteristics and (iii) the growth and aging population over time.

In the analysis of demographic heterogeneity (Section 6) we compute 4 different outcomes for each age-class and sex as follow:

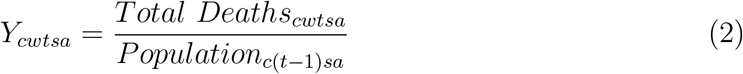

Now, *Y*_*cwtsa*_ represents the deaths per population in canton *c*, in week *w* and year *t*, for a specific gender *s* and age cohort *a*. We report descriptive statistics of the main variables in Table (1). In the Appendix (table A1) we also report the summary statistics for the same variables at the cantonal level.

**Table 1:** Descriptive Statistics by Sex and Age-class

| Variable | Male | Female | Total |
| --- | --- | --- | --- |
| Deaths | 22.56<br>(23.75) | 24.08<br>(26.13) | 46.64<br>(49.37) |
| Population | 1.48<br>(1.54) | 1.53<br>(1.59) | 3.01<br>(3.13) |
| Share Deaths | 15.62<br>(6.40) | 16.06<br>(6.84) | 15.84<br>(5.07) |
| Deaths (0-29) | 0.56<br>(0.96) | 0.30<br>(0.66) | 0.86<br>(1.33) |
| Population (0-29) | 0.52<br>(0.53) | 0.50<br>(0.51) | 1.01<br>(1.04) |
| Share Deaths (0-29) | 1.09<br>(2.64) | 0.60<br>(2.02) | 0.85<br>(1.70) |
| Deaths (30-59) | 2.54<br>(3.08) | 1.48<br>(1.97) | 4.03<br>(4.63) |
| Population (30-59) | 0.66<br>(0.70) | 0.66<br>(0.70) | 1.32<br>(1.40) |
| Share Deaths (30-59) | 3.91<br>(4.44) | 2.31<br>(3.46) | 3.12<br>(2.85) |
| Deaths (60-90) | 16.03<br>(16.87) | 14.54<br>(15.81) | 30.58<br>(32.18) |
| Population (60-90) | 0.29<br>(0.31) | 0.36<br>(0.38) | 0.65<br>(0.68) |
| Share Deaths (60-90) | 55.86<br>(26.97) | 41.54<br>(21.68) | 48.02<br>(17.92) |
| Deaths (90+) | 3.42<br>(4.30) | 7.75<br>(9.08) | 11.17<br>(12.95) |
| Population (90+) | 0.01<br>(0.01) | 0.02<br>(0.02) | 0.02<br>(0.03) |
| Share Deaths (90+) | 561.70<br>(602.50) | 449.30<br>(322.50) | 475.60<br>(287.90) |
| Observations | 21840 |  |  |

Note: Summary statistics of the main variables for Switzerland. Each row contains the mean of the variable for male, female and total population. Standard deviation in parenthesis. Observations contain (i) weekly data of deaths for the first 40 weeks of each year between 2000 and 2020 (ii) yearly data of population between 1999 and 2019. *Share Deaths* is the ratio between total deaths in year $t$ and population in $t - 1$ . Variables are expressed in 100,000 ratio.

## 4 Identification Strategy

To identify the causal effect of mandatory face-mask policy on deaths of all causes, we employ a difference-in-difference model with fixed-effects and an event study. It is worth to recall three important dates of our Policy Timeline. First, on July 6, the Federal Council imposes face-masks wearing on public transportation for the whole of Switzerland. Second, between July 7 and October 4, 9 cantons applied stricter face-mask policies extending the mandate to restaurants, shops, etc.. Finally, starting from October 18 the Federal Authorities made face-masks compulsory in all public indoor spaces. Thus, in our research design, a *Treated* canton has a mandate of mask wearing in shops, bars and restaurants while a *Control* canton has compulsory mask wearing on public transport only. Hence, we estimate the effect of such additional requirements.

### 4.1 Difference-in-Difference

We estimate the following equation:

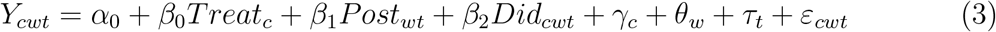

With *Treat* being a dummy identifying whether the cantons adopted stricter face-mask policy or not; *Post* is a dummy taking value 1 in the post-policy period, 0 otherwise. Then, *Did* is the interaction between *Treat* and *Post*. Our parameter of interest is *β*_2_. Finally, *γ*_*c*_, *θ*_*w*_ and *τ*_*t*_ are respectively canton, week and year fixed-effects. Table (2) reports the estimates of equation (3). Importantly, we set a unique starting date for the post-period for all cantons on July 7. This allows us to get rid off likely anticipation bias due to behavioral responses. For example, people in cantons that would implement the policy at a later date may decide to wear a face-mask in indoor places irrespective to canton’s measures and this would be particularly so in cantons that would later activate the same policy. However, attributing the treatment earlier then the actual date of adoption in some cantons might lead to downward estimated coefficient. In practice, to an extreme this would consider as treated some cantons that would be controls at that point. Thus, we test whether this attenuation is relevant in the event study, and in a staggered difference-in-difference, where the actual date of implementation is exploited (see tables (A6)-(A10) in Appendix A).

**Table 2:** Difference-in-Difference regression on share deaths

| VARIABLES | (1)<br>Male | (2)<br>Male | (3)<br>Male | (4)<br>Female | (5)<br>Female | (6)<br>Female | (7)<br>Total | (8)<br>Total | (9)<br>Total |
| --- | --- | --- | --- | --- | --- | --- | --- | --- | --- |
| $Treat^{(a)}$ | 0.525<br>(0.822) | 0.544***<br>(0.009) | 0.544***<br>(0.009) | 0.511<br>(1.119) | 1.589***<br>(0.012) | 1.589***<br>(0.012) | 0.527<br>(0.960) | 1.075***<br>(0.007) | 1.075***<br>(0.00654) |
| $Post^{(b)}$ | -0.673<br>(0.476) | -0.673<br>(0.476) | 0.0950<br>(0.587) | -1.979***<br>(0.632) | -1.979***<br>(0.632) | -0.446<br>(0.619) | -1.318***<br>(0.259) | -1.318***<br>(0.259) | -0.161<br>(0.330) |
| DiD | -1.167*<br>(0.600) | -1.167*<br>(0.601) | -1.167*<br>(0.602) | 0.357<br>(0.789) | 0.357<br>(0.790) | 0.357<br>(0.791) | -0.414<br>(0.422) | -0.414<br>(0.422) | -0.414<br>(0.423) |
| Constant | 15.45***<br>(0.421) | 13.79***<br>(0.007) | 16.75***<br>(0.307) | 15.91***<br>(0.499) | 13.90***<br>(0.010) | 16.66***<br>(0.465) | 15.68***<br>(0.458) | 13.85***<br>(0.004) | 16.71***<br>(0.265) |
| Observations | 21,840 | 21,840 | 21,840 | 21,840 | 21,840 | 21,840 | 21,840 | 21,840 | 21,840 |
| R-squared | 0.002 | 0.085 | 0.121 | 0.002 | 0.122 | 0.188 | 0.004 | 0.172 | 0.255 |
| Year FE | NO | NO | YES | NO | NO | YES | NO | NO | YES |
| Canton FE | NO | YES | YES | NO | YES | YES | NO | YES | YES |
| Week FE | NO | NO | YES | NO | NO | YES | NO | NO | YES |
| Mean | 15.62 | 15.62 | 15.62 | 16.06 | 16.06 | 16.06 | 15.84 | 15.84 | 15.84 |

Note: Results of regression in Equation (3). Dependent variable defined as equation (1): ratio between total deaths of a specific canton in a week and year $t$ on total population of the canton in year $t-1$ . Column 1-2-3 contain observation for males. Columns 4-5-6 contain observations for females. Column 7-8-9 contain observations for aggregate male and female population. S.E. clustered at a canton level (\*\*\*) $p < 0.01$ , \*\* $p < 0.05$ , \* $p < 0.1$ . Period of analysis: between January 2000 and October, 4th 2020. (a) $Treated$ cantons are those that between July, 7 and October, 4 imposed any mask requirement other than Federal indications (e.g in supermarket, restaurants, open space): BS, FR, GE, JU, NE, SO, VS, VD, ZH. (b) $Post$ is equal to 1 for all cantons after July, 7.

With the main identification we find a marginally significant (10% level) reduction in total mortality only for males (−1.17): this corresponds to a reduction of 7.5% in weekly deaths. While for female we find a statistically insignificant increase in mortality (+0.36, corresponding to a 2.2% increase in weekly share deaths). When looking at total deaths (columns 7-9) the coefficient is -0.41, and not significant. Hence, it appears that the impact of the policy is heterogeneous across genders. In Section 6 we dig deeper in the demographic heterogeneity.

### 4.2 Event Study

In this section we estimate the panel event study keeping the same outcomes as in the previous section, but allowing the date of the event to be the actual date of implementation of the policy, and therefore it varies by canton. The policy lags actually allow to accommodate the temporal nature of treatment effects. Hence, with the full set of event lags we can inspect parallel trends in the pre-treatment period (Clarke and Schythe, 2020). Formally, we estimate the following equation:

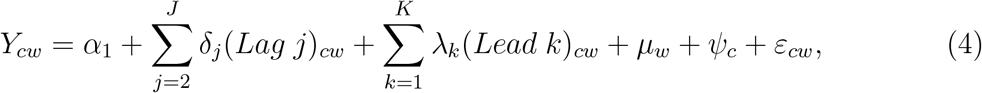

Where *ψ*_*c*_ and *µ*_*w*_ are canton and week fixed effects and *ε*_*cw*_ is an unobserved error term. Further *Lag*_*j*_ and *Lead*_*k*_ are two binary variables indicating that canton *c* is *j /k* weeks far from mask requirement. Formally, we define *Lag*_*j*_ and *Lead*_*k*_ according to equations(5)-(8)

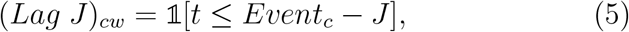

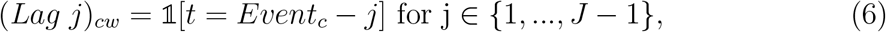

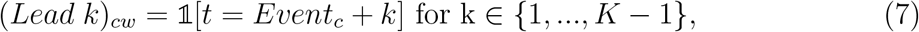

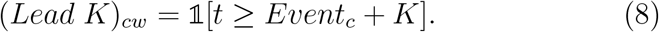

Where, *Event*_*c*_ is a variable indicating the week *w* in which the face mask requirement is adopted in canton *c*. The first *Lag* is omitted to capture the baseline difference between treated and control cantons. In figures (3)-(5) we provide a graphical representation of the estimates. Blue dots indicate the point estimates, while the light blue area represents the confidence interval at the 99% level. The event study approach confirms the validity of our design as in the pre-period (to the left of the vertical line) we have no significant differences. At the same time, the event study approach casts some doubt on the robustness of the previous results. Indeed, Figures 3, 4, and 5 show no significant effects. In fact if anything it seems that the last point estimates are rather on the positive side, i.e. increase mortality. In Figures (A1)-(A3) in Appendix A, we implement the event study approach on the four age groups and by gender, we again find no significant effects of the stricter mask-wearing requirements.

**Figure 3:**
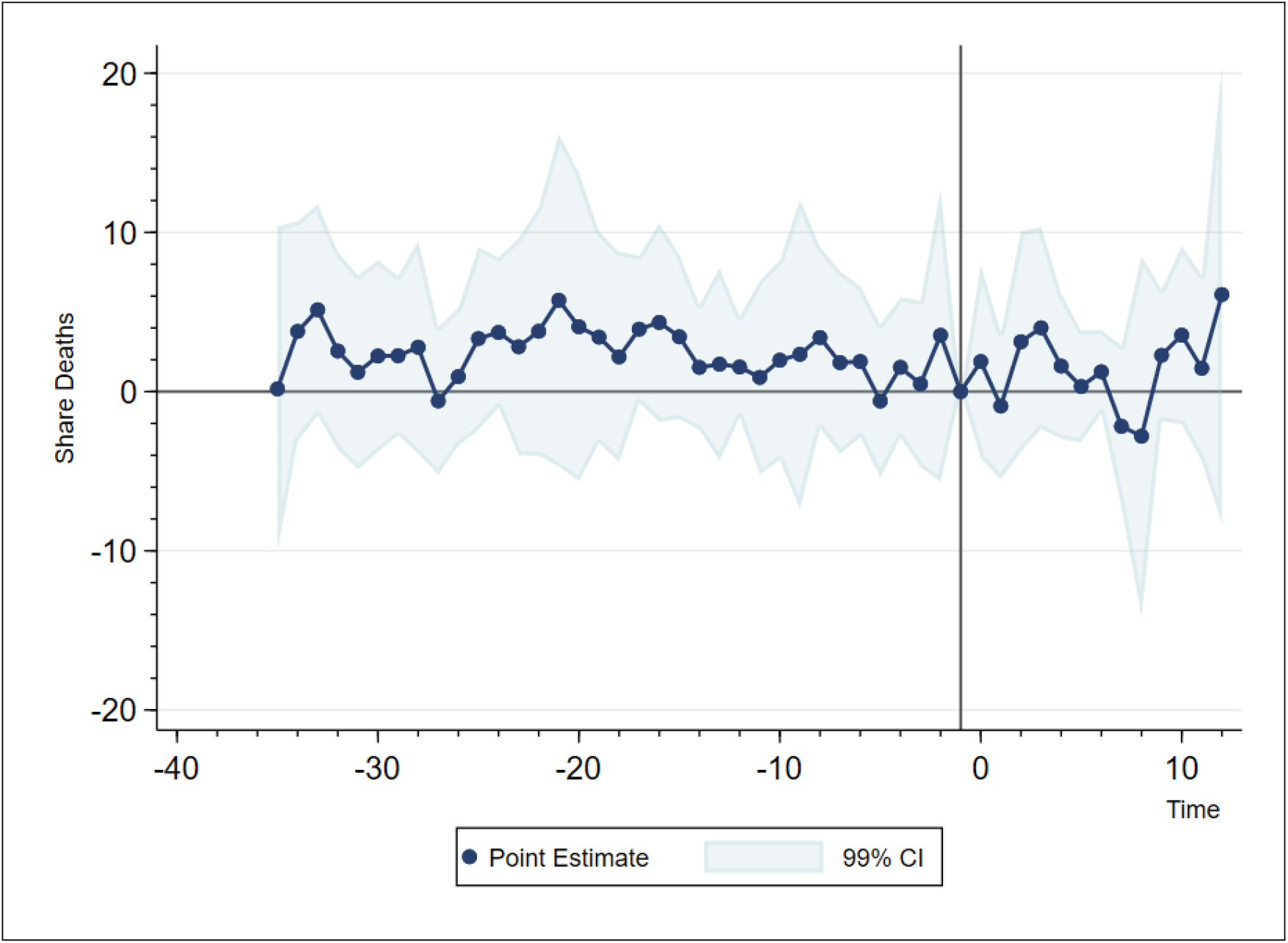
Event study. Male Deaths Notes: Estimates of equation (4). Point estimates (blue points) are displayed along with their 99% confidence intervals (light blue area). Baseline period: 1 week prior to the adoption of the face masks policy in each adopting canton, indicated by the solid vertical line in the plot.

**Figure 4:**
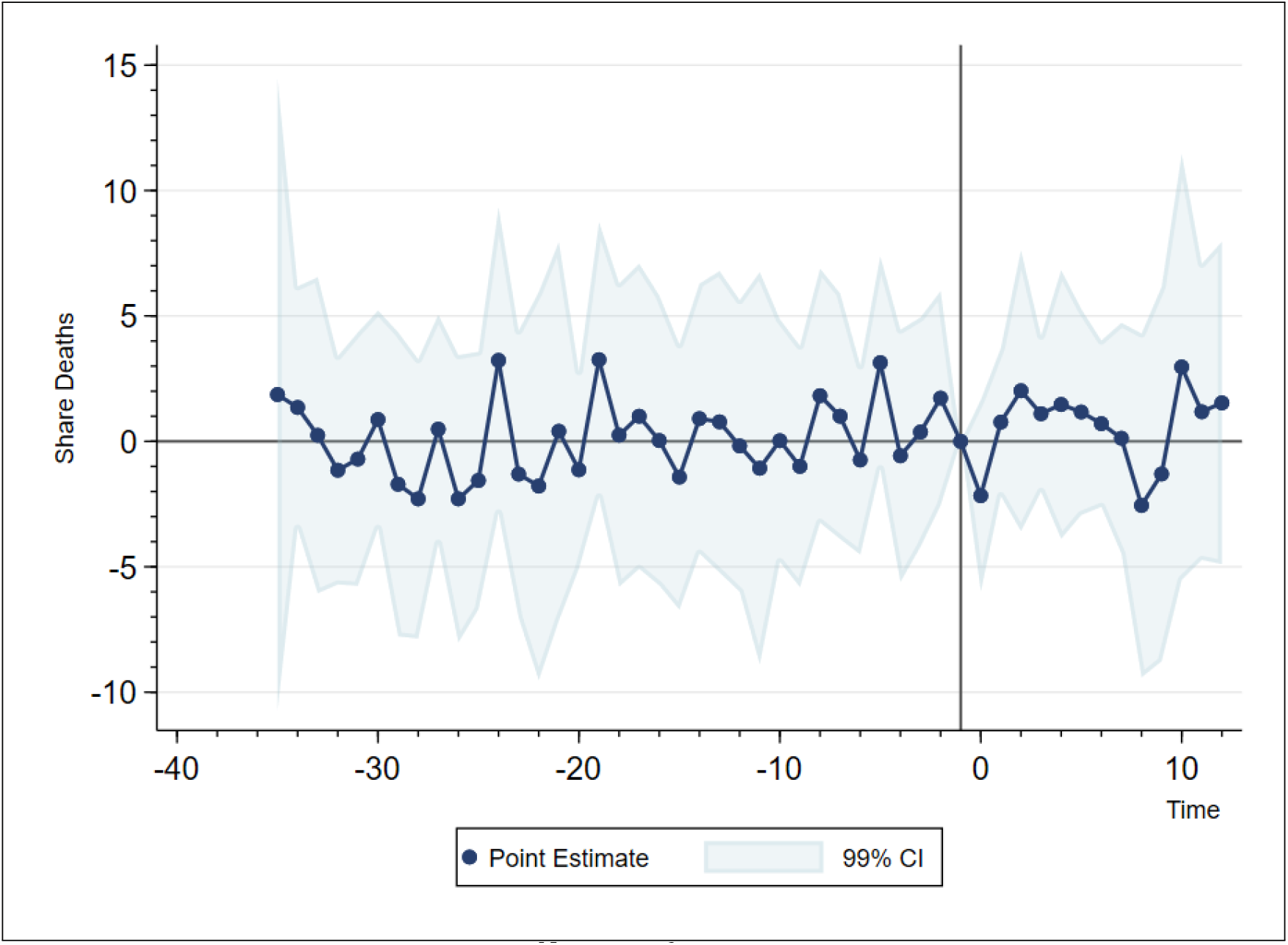
Event study. Female Deaths Notes: see figure 3

**Figure 5:**
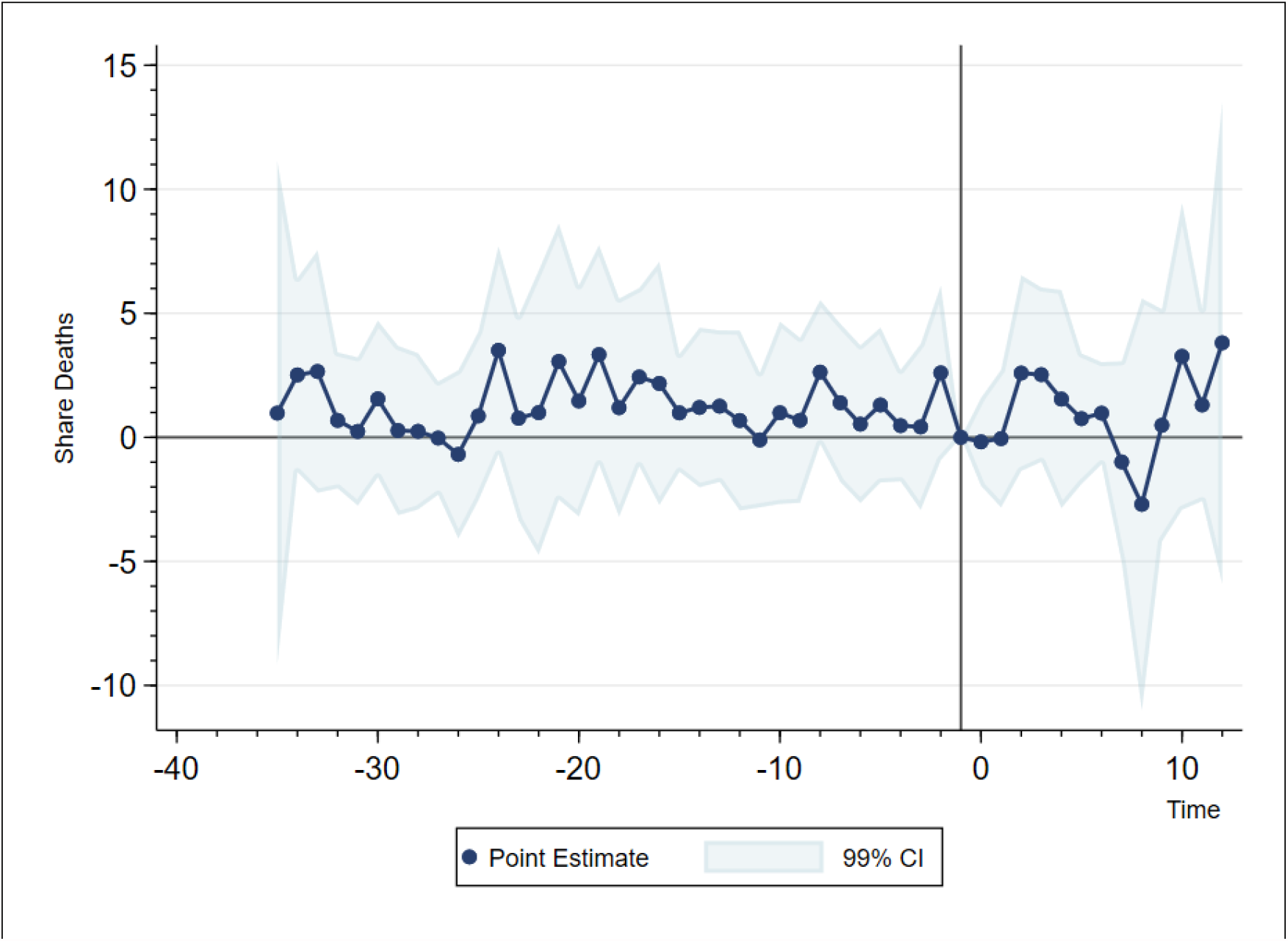
Event study. Deaths Total Population Notes: see figure 3

## 5 Robustness checks and extra results

In this section we provide some analysis of robustness.

### 1. Pre-trends

The event study approach in the previous section shows that parallel trends assumption seems to hold.

### 2. Weighted Analysis

Following Solon et al., 2015, we provide the results of the weighted regression. In particular, we estimate our baseline model (equation 3) using cantonal population size as analytic weights (table 3).

**Table 3:** Weighted Difference-in-Difference on share deaths

| VARIABLES | (1)<br>Male | (2)<br>Male | (3)<br>Male | (4)<br>Female | (5)<br>Female | (6)<br>Female | (7)<br>Total | (8)<br>Total | (9)<br>Total |
| --- | --- | --- | --- | --- | --- | --- | --- | --- | --- |
| Treat | -0.601<br>(0.760) | 0.476***<br>(0.00589) | 0.478***<br>(0.00587) | -0.421<br>(0.871) | 1.538***<br>(0.00625) | 1.538***<br>(0.00625) | -0.507<br>(0.800) | 1.014***<br>(0.00504) | 1.015***<br>(0.00504) |
| Post | -0.893***<br>(0.281) | -0.852***<br>(0.286) | 0.102<br>(0.411) | -1.783***<br>(0.273) | -1.739***<br>(0.276) | -0.148<br>(0.337) | -1.342***<br>(0.244) | -1.299***<br>(0.249) | -0.0199<br>(0.350) |
| DiD | -0.832**<br>(0.336) | -0.832**<br>(0.333) | -0.842**<br>(0.332) | -0.119<br>(0.363) | -0.104<br>(0.359) | -0.109<br>(0.359) | -0.475<br>(0.293) | -0.469<br>(0.288) | -0.476<br>(0.287) |
| Constant | 15.54***<br>(0.600) | 13.80***<br>(0.00504) | 16.70***<br>(0.226) | 15.99***<br>(0.669) | 13.90***<br>(0.00480) | 17.24***<br>(0.307) | 15.76***<br>(0.634) | 13.85***<br>(0.00436) | 16.97***<br>(0.230) |
| Observations | 21,840 | 21,840 | 21,840 | 21,840 | 21,840 | 21,840 | 21,840 | 21,840 | 21,840 |
| R-squared | 0.008 | 0.172 | 0.262 | 0.005 | 0.216 | 0.356 | 0.009 | 0.279 | 0.446 |
| Year FE | NO | NO | YES | NO | NO | YES | NO | NO | YES |
| Canton FE | NO | YES | YES | NO | YES | YES | NO | YES | YES |
| Week FE | NO | NO | YES | NO | NO | YES | NO | NO | YES |
| Mean | 15.22 | 15.22 | 15.22 | 15.75 | 15.75 | 15.75 | 15.49 | 15.49 | 15.49 |

Note: Results of regression in Equation (3) weighted using population as analytical weights. Column 1-2-3 contain observation for male population. Columns 4-5-6 contain observations for female population. Column 7-8-9 contain observations for aggregate male and female population. S.E. clustered at a canton level (\*\*\*) $p < 0.01$ , \*\* $p < 0.05$ , \* $p < 0.1$ ). Period of estimation: between January 2000 and October, 4th 2020.
(a) *Treated* cantons are those that between July, 7 and October, 4 have imposed any mask requirement other then Federal indications (e.g in supermarket, restaurants, open space): BS, FR, GE, JU, NE, SO, VS, VD, ZH.
(b) *Post* is equal to 1 for all cantons after July, 7.

It is easy to see that the estimated effects are in line with those presented earlier, if anything the effect for males is slightly smaller at −.842, a 5.5% reduction in mortality, but more precisely estimated *p* − *value <* .05.

### 3. Dynamic Betas

In our main identification we assume *β* to be constant over all the weeks after the implementation of the stricter requirements. However, policy effects might vary as time goes by. Accordingly, in this section we estimate a dynamic model where *β* can vary across weeks (this model is similar in spirit to the event study approach). Formally, we estimate the following equation.

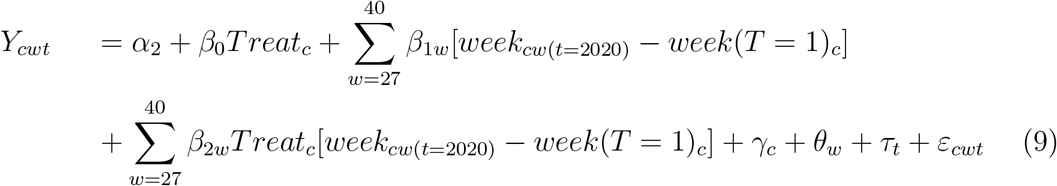

In particular, *Treat*_*c*_ is a dummy equal to 1 if canton *c* is ever treated. Then, [*week* − *week*(*T* = 1)_*c*_] is the difference between the observation week and the first week of implementation of the extra measure in canton *c*. The parameters of interest are the *β*_2*w*_ and represent the mean difference in the outcome of interest in a specific week *w*. We also control with canton (*γ*_*c*_), week (*θ*_*w*_) and year (*τ*_*t*_) fixed-effects. In figures (6)-(8) we plot the dynamic coefficient (*β*_2*w*_). The post-policy begins in week 26 to on (July 7) for all cantons. For each week after the treatment we report the point estimates and their standard error. Figures (6)-(8) confirm the overall modest achievement of the policy. The heterogeneity of results across sex is confirmed as well. Figure (6) reports the coefficients for males. After two weeks of treatment there is a sharp and significant decrease in the estimated coefficient. That very positive outcome is however likely due to sample variability as it would be too soon for the policy to produce such a sharp effect and also it appears as an isolated drop as we can see in later weeks. Then, between week 3 and 9 the points estimates lie below the 0-line. After a further increase in week 10, the outcome mean is again reduced between weeks 11 and 12. Figure (7) reports the weekly coefficients for females. In the first three weeks after the introduction of the policy there is a drop in the coefficients. However, in the subsequent weeks the points lie mostly on the positive side of the graphs. In Appendix A, Figures (A4)-(A6) show dynamic *β*_2*w*_ for the four age cohorts and by gender. On a quick inspection it is clear that the effect in week 2 is driven by the very “unusual” decline in mortality among men aged 60-90. In brief, we confirm that the effects are at best very modest across age, gender, and over different time horizons.

**Figure 6:**
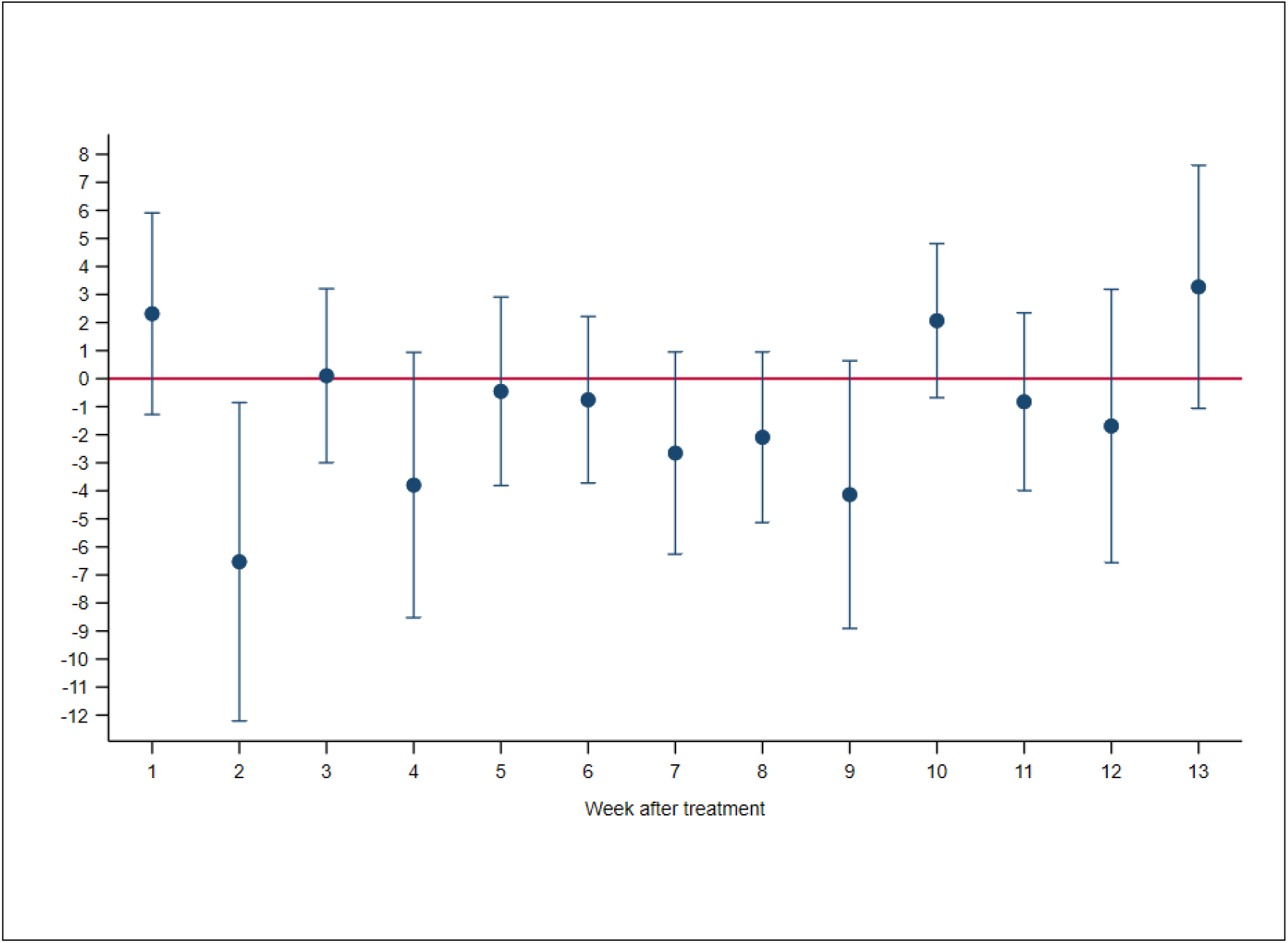
Difference-in-Difference regression on male deaths with dynamic Beta Note: Blue dots are the estimated *β*_2_ in week *w* as in equation 9. Week 1 is the first week after the treatment, until the 13^*t*^*h* week. Outcome defined as 1. Each bar represents the clustered SE of the point estimates (*** p*<*0.01, ** p*<*0.05, * p*<*0.1).

**Figure 7:**
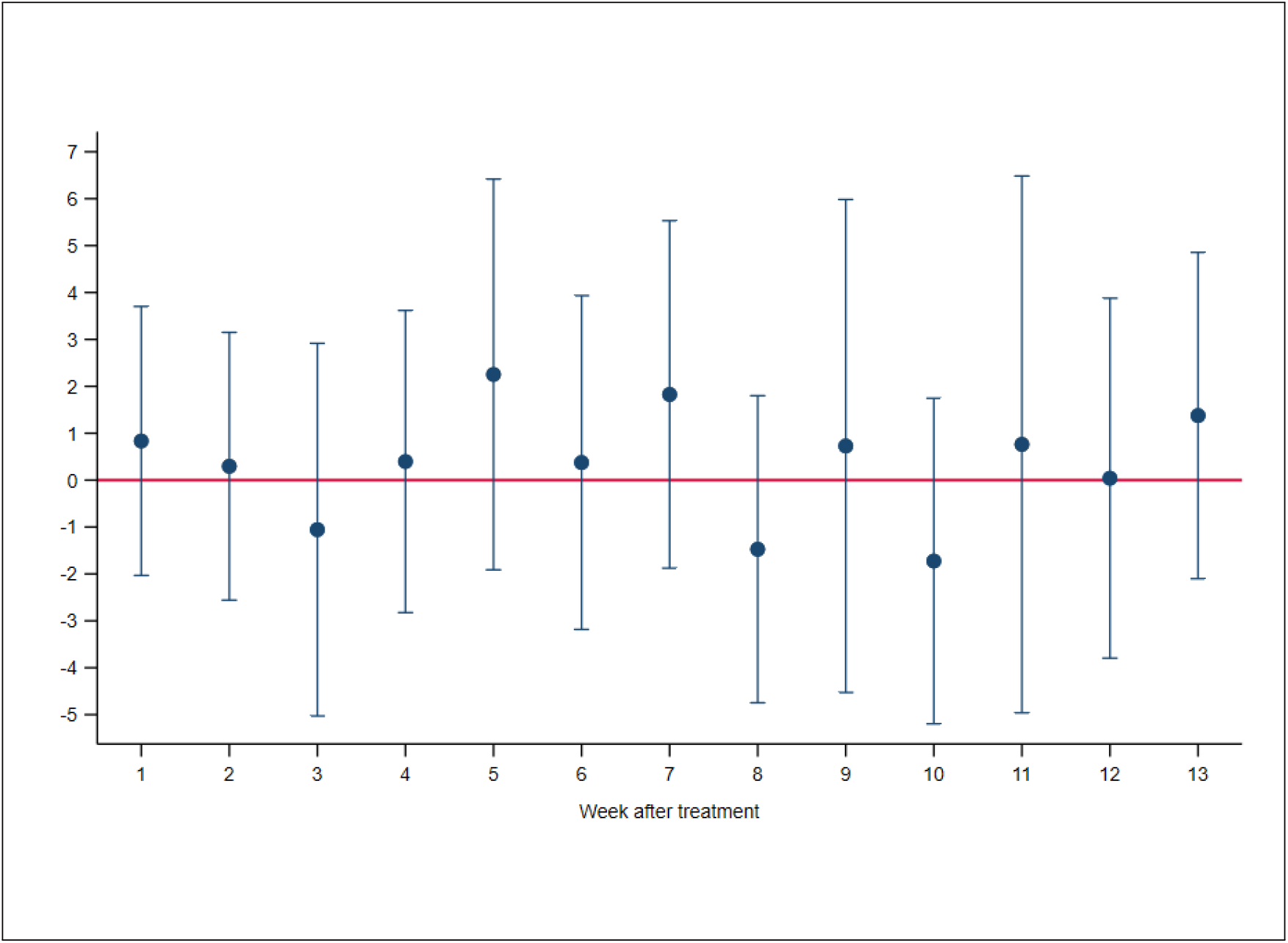
Difference-in-Difference regression on female deaths with dynamic Beta Note: see figure 6

**Figure 8:**
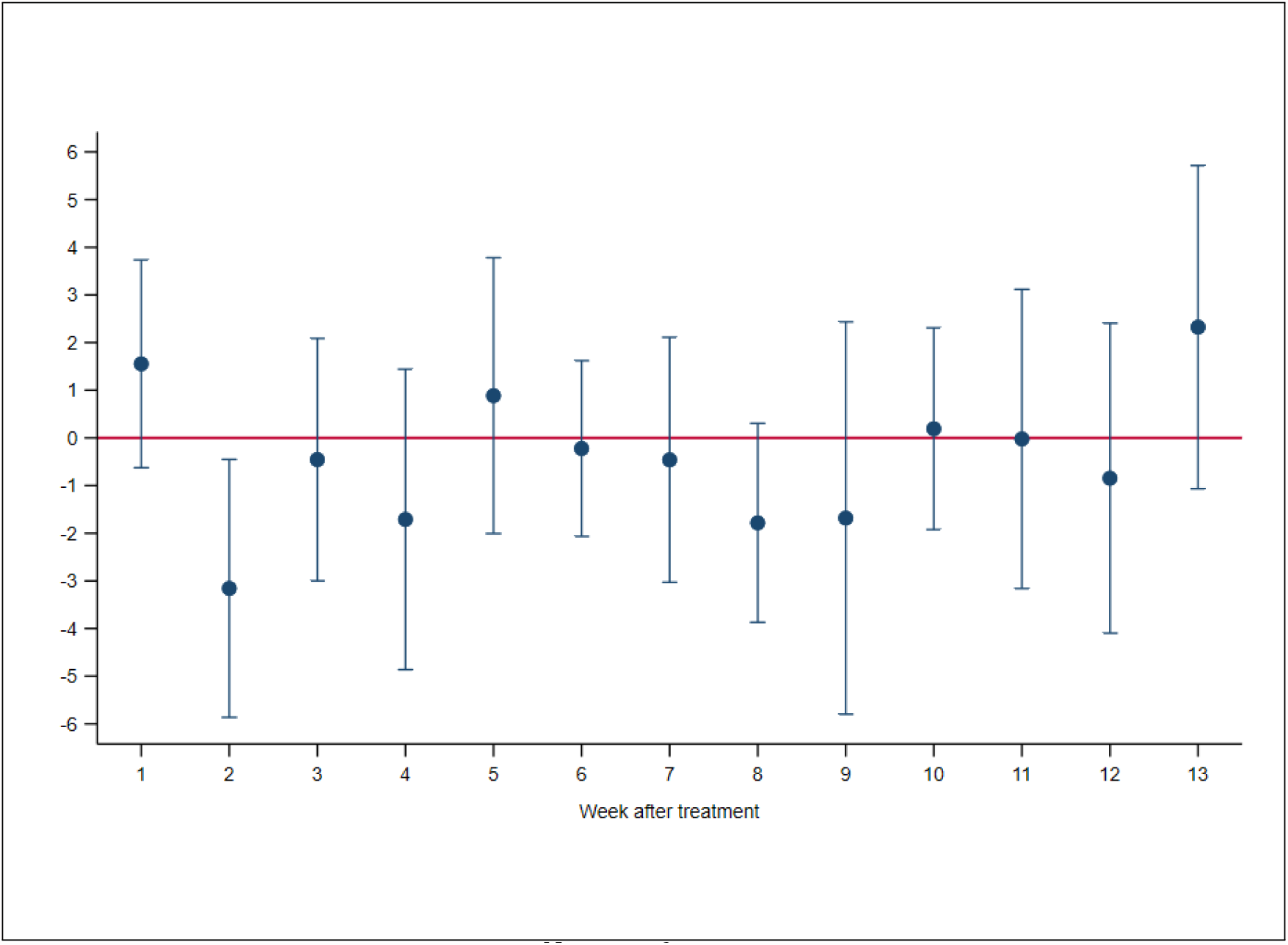
Difference-in-Difference regression on total deaths with dynamic Beta Note: see figure 6

## 6 Heterogeneity analysis

In this section we explore the likely mechanisms driving our findings. First, we study the heterogeneous impact of the policy on 4 different age classes. Second, we conduct a policy-mix analysis in which mandatory face-mask is combined with other NPIs.

### 6.1 Demographic analysis

In the demographic analysis we estimate the effect of face-mask policy on different age-classes. Accordingly, we group the share of deaths in four cohorts: young (0-29), middle (30-59), old (60-90) and very old (over 90). We choose the age brackets according to WHO standards. We first estimate our Difference-in-Difference model (in equation 3) in each age group, controlling with time and canton fixed-effects. Then, we employ a pooled regression.

Estimating the effect on each age-class separately requires cohort-specific pre-trend periods. Indeed, for 30-59 class we can keep the whole pre-period. While, for the three other cohorts we restrict the sample according to the periods in which the outcome of treatment and control groups show parallel trends.^4^ In tables (4)-(7) the estimates are reported.

**Table 4:** Difference-in-Difference regression on share deaths. Age-class: 0-29

| VARIABLES | (1)<br>Male<br>(0-29) | (2)<br>Male<br>(0-29) | (3)<br>Male<br>(0-29) | (4)<br>Female<br>(0-29) | (5)<br>Female<br>(0-29) | (6)<br>Female<br>(0-29) | (7)<br>Total<br>(0-29) | (8)<br>Total<br>(0-29) | (9)<br>Total<br>(0-29) |
| --- | --- | --- | --- | --- | --- | --- | --- | --- | --- |
| *Treat*<sup>(a)</sup> | -0.045<br>(0.045) | -0.005<br>(0.006) | -0.005<br>(0.006) | -0.004<br>(0.037) | 0.083***<br>(0.005) | 0.083***<br>(0.005) | -0.028<br>(0.037) | 0.037***<br>(0.004) | 0.037***<br>(0.004) |
| *Post*<sup>(b)</sup> | -0.010<br>(0.165) | -0.010<br>(0.166) | 0.022<br>(0.234) | -0.030<br>(0.195) | -0.030<br>(0.195) | 0.022<br>(0.237) | -0.020<br>(0.152) | -0.020<br>(0.152) | 0.020<br>(0.183) |
| DiD | 0.061<br>(0.235) | 0.061<br>(0.235) | 0.061<br>(0.236) | 0.010<br>(0.209) | 0.010<br>(0.209) | 0.010<br>(0.209) | 0.036<br>(0.163) | 0.036<br>(0.163) | 0.036<br>(0.163) |
| Constant | 0.968***<br>(0.031) | 0.923***<br>(0.004) | 1.079***<br>(0.128) | 0.543***<br>(0.021) | 0.546***<br>(0.005) | 0.571***<br>(0.105) | 0.762***<br>(0.025) | 0.739***<br>(0.004) | 0.831***<br>(0.056) |
| Observations | 13,520 | 13,520 | 13,520 | 13,520 | 13,520 | 13,520 | 13,520 | 13,520 | 13,520 |
| R-squared | 0.000 | 0.002 | 0.007 | 0.000 | 0.002 | 0.006 | 0.000 | 0.003 | 0.007 |
| Year FE | NO | NO | YES | NO | NO | YES | NO | NO | YES |
| Canton FE | NO | YES | YES | NO | YES | YES | NO | YES | YES |
| Week FE | NO | NO | YES | NO | NO | YES | NO | NO | YES |
| Mean | 0.952 | 0.952 | 0.952 | 0.541 | 0.541 | 0.541 | 0.752 | 0.752 | 0.752 |

Note: Period of estimation is between January 2008 and October, 4th 2020. Results of regression in Equation (3). Dependent variable defined as equation (2): ratio between deaths in age-class *age* of a specific canton in a week and year *t* on total population of age *age* in the canton in year *t-1*. Column 1-2-3 contain observation for male population. Columns 4-5-6 contain observations for female population. Column 7-8-9 contain observations for aggregate male and female population. S.E. clustered at a canton level (\*\*\*) $p < 0.01$ , \*\* $p < 0.05$ , \* $p < 0.1$ ).
(a) *Treated* cantons are those that between July, 7 and October, 4 have imposed any mask requirement other then Federal indications (e.g in supermarket, restaurants, open space): BS, FR, GE, JU, NE, SO, VS, VD, ZH.
(b) *Post* is equal to 1 for all cantons after July, 7.

**Table 5:** Difference-in-Difference regression on share deaths. Age-class: 30-59

| VARIABLES | (1)<br>Male<br>(30-59) | (2)<br>Male<br>(30-59) | (3)<br>Male<br>(30-59) | (4)<br>Female<br>(30-59) | (5)<br>Female<br>(30-59) | (6)<br>Female<br>(30-59) | (7)<br>Total<br>(30-59) | (8)<br>Total<br>(30-59) | (9)<br>Total<br>(30-59) |
| --- | --- | --- | --- | --- | --- | --- | --- | --- | --- |
| *Treat<sup>(a)</sup>* | 0.515***<br>(0.178) | -0.017***<br>(0.005) | -0.017***<br>(0.005) | 0.183*<br>(0.105) | -0.084***<br>(0.005) | -0.084***<br>(0.005) | 0.337**<br>(0.138) | -0.051***<br>(0.004) | -0.051***<br>(0.004) |
| *Post<sup>(b)</sup>* | -1.119***<br>(0.270) | -1.119***<br>(0.270) | -0.233<br>(0.279) | -0.789***<br>(0.202) | -0.789***<br>(0.202) | -0.288<br>(0.240) | -0.956***<br>(0.193) | -0.956***<br>(0.194) | -0.261<br>(0.235) |
| DiD | 0.127<br>(0.312) | 0.127<br>(0.312) | 0.127<br>(0.313) | 0.340<br>(0.318) | 0.340<br>(0.318) | 0.340<br>(0.319) | 0.235<br>(0.274) | 0.235<br>(0.274) | 0.235<br>(0.274) |
| Constant | 3.743***<br>(0.097) | 3.634***<br>(0.004) | 5.133***<br>(0.300) | 2.254***<br>(0.062) | 2.293***<br>(0.003) | 2.709***<br>(0.209) | 3.012***<br>(0.075) | 2.972***<br>(0.003) | 3.944***<br>(0.156) |
| Observations | 21,840 | 21,840 | 21,840 | 21,840 | 21,840 | 21,840 | 21,840 | 21,840 | 21,840 |
| R-squared | 0.004 | 0.013 | 0.029 | 0.001 | 0.007 | 0.015 | 0.005 | 0.017 | 0.038 |
| Year FE | NO | NO | YES | NO | NO | YES | NO | NO | YES |
| Canton FE | NO | YES | YES | NO | YES | YES | NO | YES | YES |
| Week FE | NO | NO | YES | NO | NO | YES | NO | NO | YES |
| Mean | 3.905 | 3.905 | 3.905 | 2.307 | 2.307 | 2.307 | 3.115 | 3.115 | 3.115 |

Note: Period of estimation is between January 2000 and October, 4th 2020. See table 4 for other details.

**Table 6:** Difference-in-Difference regression on share deaths. Age-class: 60-90

| VARIABLES | (1)<br>Male<br>(60-90) | (2)<br>Male<br>(60-90) | (3)<br>Male<br>(60-90) | (4)<br>Female<br>(60-90) | (5)<br>Female<br>(60-90) | (6)<br>Female<br>(60-90) | (7)<br>Total<br>(60-90) | (8)<br>Total<br>(60-90) | (9)<br>Total<br>(60-90) |
| --- | --- | --- | --- | --- | --- | --- | --- | --- | --- |
| *Treat<sup>(a)</sup>* | 3.595**<br>(1.597) | 1.209***<br>(0.064) | 1.209***<br>(0.064) | 0.889<br>(1.440) | 1.417***<br>(0.055) | 1.417***<br>(0.055) | 1.921<br>(1.422) | 1.109***<br>(0.031) | 1.109***<br>(0.031) |
| *Post<sup>(b)</sup>* | -5.946***<br>(1.438) | -5.946***<br>(1.440) | -0.984<br>(1.769) | -6.264***<br>(1.306) | -6.264***<br>(1.308) | -0.417<br>(1.173) | -6.012***<br>(0.698) | -6.012***<br>(0.699) | -0.667<br>(0.944) |
| DiD | -0.677<br>(1.773) | -0.677<br>(1.775) | -0.677<br>(1.779) | 2.403<br>(1.509) | 2.403<br>(1.511) | 2.403<br>(1.515) | 0.987<br>(0.862) | 0.987<br>(0.863) | 0.987<br>(0.865) |
| Constant | 48.502***<br>(0.847) | 47.700***<br>(0.052) | 53.138***<br>(1.529) | 37.689***<br>(0.808) | 36.463***<br>(0.047) | 42.148***<br>(1.569) | 42.799***<br>(0.773) | 41.788***<br>(0.025) | 47.203***<br>(0.961) |
| Observations | 9,360 | 9,360 | 9,360 | 9,360 | 9,360 | 9,360 | 9,360 | 9,360 | 9,360 |
| R-squared | 0.008 | 0.034 | 0.069 | 0.004 | 0.034 | 0.076 | 0.009 | 0.053 | 0.117 |
| Year FE | NO | NO | YES | NO | NO | YES | NO | NO | YES |
| Canton FE | NO | YES | YES | NO | YES | YES | NO | YES | YES |
| Week FE | NO | NO | YES | NO | NO | YES | NO | NO | YES |
| Mean | 49.524 | 49.524 | 49.524 | 37.800 | 37.800 | 37.800 | 43.259 | 43.259 | 43.259 |

Note: Period of estimation is between January 2012 and October, 4th 2020. See table 4 for other details.

**Table 7:** Difference-in-Difference regression on share deaths. Age-class: 90+

| VARIABLES | (1)<br>Male<br>(90+) | (2)<br>Male<br>(90+) | (3)<br>Male<br>(90+) | (4)<br>Female<br>(90+) | (5)<br>Female<br>(90+) | (6)<br>Female<br>(90+) | (7)<br>Total<br>(90+) | (8)<br>Total<br>(90+) | (9)<br>Total<br>(90+) |
| --- | --- | --- | --- | --- | --- | --- | --- | --- | --- |
| *Treat*<sup>(a)</sup> | -24.88**<br>(11.62) | -27.48***<br>(1.33) | -27.48***<br>(1.33) | -29.18**<br>(10.64) | -29.41***<br>(1.01) | -29.41***<br>(1.02) | -26.81**<br>(10.08) | -29.84***<br>(0.93) | -29.84***<br>(0.93) |
| *Post*<sup>(b)</sup> | 33.49<br>(43.37) | 33.49<br>(43.40) | 125.67**<br>(49.35) | -23.78<br>(15.58) | -23.78<br>(15.59) | 47.74**<br>(20.94) | -5.63<br>(19.27) | -5.63<br>(19.28) | 70.08***<br>(23.60) |
| DiD | -29.08<br>(47.04) | -29.08<br>(47.07) | -29.08<br>(47.15) | 35.47<br>(35.86) | 35.47<br>(35.89) | 35.47<br>(35.95) | 17.66<br>(32.83) | 17.66<br>(32.85) | 17.66<br>(32.91) |
| Constant | 553.77***<br>(9.32) | 546.00***<br>(1.23) | 664.62***<br>(30.54) | 452.68***<br>(6.50) | 464.85***<br>(0.44) | 554.30***<br>(13.80) | 476.37***<br>(6.28) | 488.31***<br>(0.55) | 584.47***<br>(13.31) |
| Observations | 18,144 | 18,144 | 18,144 | 18,144 | 18,144 | 18,144 | 18,144 | 18,144 | 18,144 |
| R-squared | 0.00 | 0.01 | 0.03 | 0.00 | 0.01 | 0.06 | 0.00 | 0.01 | 0.09 |
| Year FE | NO | NO | YES | NO | NO | YES | NO | NO | YES |
| Canton FE | NO | YES | YES | NO | YES | YES | NO | YES | YES |
| Week FE | NO | NO | YES | NO | NO | YES | NO | NO | YES |
| Mean | 546.15 | 546.15 | 546.15 | 442.62 | 442.62 | 442.62 | 467.44 | 467.44 | 467.44 |

Note: Period of estimation is between January 2008 and October, 4th 2020. See table 4 for other details.

Results in tables (4)-(7) are not statistical significant in any of the age-class. Table (4) reports a small increase in young mortality for both sexes (+6.45% male and +1.84% female). Also Table (5) shows an increase in the mean outcome for both genders: +0.13 male and +0.34 female, corresponding respectively to an increase of 3.25% and 14.7% in weekly mortality. Within the older cohorts (tables 6-7) the effects of the policy are heterogeneous with respect to the sex. For old population (60-90) the estimated coefficients are negative for male (−0.677) indicating a reduction of 1.36% in share deaths; while for female are positive (+2.403) showing an increase of 6.3% in share deaths. For oldest population (90+) we find a decrease of -16.62% in weakly male deaths; while there is an increase +5.96% in weakly female deaths. Even not significant from a statistical point of view, the magnitude of these findings is consistent with the significant reduction in male share deaths we find in the main identification (table 2). Thus, we further investigate the heterogeneous impact of the policy with a pooled regression. The pooled regression allows us to explore the contribute of each age-group to the aggregate estimate. In particular, we use the young age-class (0-29) as baseline and estimate the partial effect for the three other cohorts. The intervention causes a barely significant reduction (at the 10% level) of -95.5 in male share deaths within 90+ age-class (Table 8). This is the partial reduction in 90+ male mortality mean with respect to the mean of deaths in young class. On the other hand, for female we find a partial increase in the mean outcome for the three age-classes, with significant results at the 10% level (+2.88) in 60-90 cohort (Table 9). When we consider total population we do not find any significant effect for the three age cohorts. Hence, we conclude that on old male the policy has some positive effect, while on female the effect is null or negative at all. Further, we prove that the small effect found for male population is mostly driven by a decrease in 90+ population’s mortality. Table (A2) - in Appendix A - also show the estimates for a pooled-regression on gender.

**Table 8:**
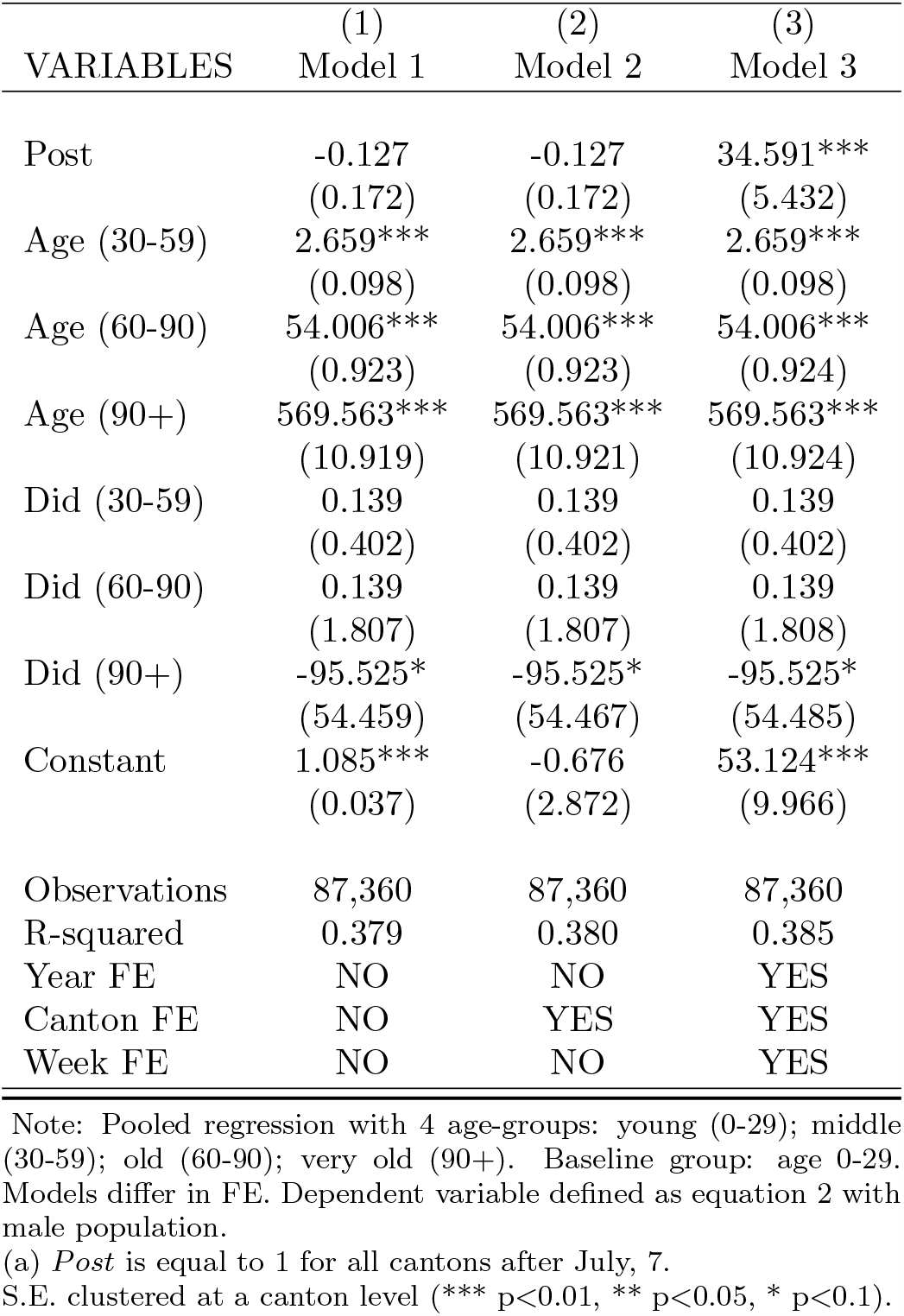
Pooled Difference-in-Difference regression on male share deaths

| VARIABLES | (1)<br>Model 1 | (2)<br>Model 2 | (3)<br>Model 3 |
| --- | --- | --- | --- |
| Post | -0.127<br>(0.172) | -0.127<br>(0.172) | 34.591***<br>(5.432) |
| Age (30-59) | 2.659***<br>(0.098) | 2.659***<br>(0.098) | 2.659***<br>(0.098) |
| Age (60-90) | 54.006***<br>(0.923) | 54.006***<br>(0.923) | 54.006***<br>(0.924) |
| Age (90+) | 569.563***<br>(10.919) | 569.563***<br>(10.921) | 569.563***<br>(10.924) |
| Did (30-59) | 0.139<br>(0.402) | 0.139<br>(0.402) | 0.139<br>(0.402) |
| Did (60-90) | 0.139<br>(1.807) | 0.139<br>(1.807) | 0.139<br>(1.808) |
| Did (90+) | -95.525*<br>(54.459) | -95.525*<br>(54.467) | -95.525*<br>(54.485) |
| Constant | 1.085***<br>(0.037) | -0.676<br>(2.872) | 53.124***<br>(9.966) |
| Observations | 87,360 | 87,360 | 87,360 |
| R-squared | 0.379 | 0.380 | 0.385 |
| Year FE | NO | NO | YES |
| Canton FE | NO | YES | YES |
| Week FE | NO | NO | YES |

Note: Pooled regression with 4 age-groups: young (0-29); middle (30-59); old (60-90); very old (90+). Baseline group: age 0-29. Models differ in FE. Dependent variable defined as equation 2 with male population.

**Table 9:** Pooled Difference-in-Difference regression on female share deaths

| VARIABLES | (1)<br>Model 1 | (2)<br>Model 2 | (3)<br>Model 3 |
| --- | --- | --- | --- |
| Post | -0.084<br>(0.192) | -0.084<br>(0.192) | 25.203***<br>(2.355) |
| Age (30-59) | 1.656***<br>(0.066) | 1.656***<br>(0.066) | 1.656***<br>(0.066) |
| Age (60-90) | 40.939***<br>(0.831) | 40.939***<br>(0.831) | 40.939***<br>(0.831) |
| Age (90+) | 457.754***<br>(6.382) | 457.754***<br>(6.383) | 457.754***<br>(6.385) |
| Did (30-59) | 0.346<br>(0.322) | 0.346<br>(0.322) | 0.346<br>(0.322) |
| Did (60-90) | 2.878*<br>(1.503) | 2.878*<br>(1.503) | 2.878*<br>(1.504) |
| Did (90+) | 17.661<br>(23.772) | 17.661<br>(23.775) | 17.661<br>(23.783) |
| Constant | 0.598***<br>(0.018) | 4.113**<br>(1.640) | 49.465***<br>(5.979) |
| Observations | 87,360 | 87,360 | 87,360 |
| R-squared | 0.578 | 0.579 | 0.586 |
| Year FE | NO | NO | YES |
| Canton FE | NO | YES | YES |
| Week FE | NO | NO | YES |

Note: Pooled regression with 4 age-groups: young (0-29); middle (30-59); old (60-90); very old (90+). Baseline group: age 0-29. Models differ in FE. Dependent variable defined as equation 2 with female population.

**Table 10:** Pooled Difference-in-Difference regression on total share deaths

| VARIABLES | (1)<br>Model 1 | (2)<br>Model 2 | (3)<br>Model 3 |
| --- | --- | --- | --- |
| Post | -0.106<br>(0.155) | -0.106<br>(0.155) | 27.034***<br>(2.743) |
| Age (30-59) | 2.164***<br>(0.079) | 2.164***<br>(0.079) | 2.164***<br>(0.079) |
| Age (60-90) | 46.890***<br>(0.850) | 46.890***<br>(0.850) | 46.890***<br>(0.850) |
| Age (90+) | 483.163***<br>(6.576) | 483.163***<br>(6.577) | 483.163***<br>(6.579) |
| Did (30-59) | 0.245<br>(0.323) | 0.245<br>(0.323) | 0.245<br>(0.323) |
| Did (60-90) | 1.646<br>(1.010) | 1.646<br>(1.010) | 1.646<br>(1.011) |
| Did (90+) | -14.946<br>(19.972) | -14.946<br>(19.975) | -14.946<br>(19.982) |
| Constant | 0.848***<br>(0.026) | 4.103**<br>(1.717) | 52.196***<br>(4.675) |
| Observations | 87,360 | 87,360 | 87,360 |
| R-squared | 0.657 | 0.658 | 0.666 |
| Year FE | NO | NO | YES |
| Canton FE | NO | YES | YES |
| Week FE | NO | NO | YES |

Note: Pooled regression with 4 age-groups: young (0-29); middle (30-59); old (60-90); very old (90+). Baseline group: age 0-29. Models differ in FE. Dependent variable defined as equation 2 at aggregate sex level.

### 6.2 Policy mix analysis

As mentioned, the treatment considered in the main analysis is heterogeneous in nature as different cantons implemented it with additional requirements. In particular, most cantons added tracing and distancing requirements. Thus, in this section we attempt at estimating the additional effects of such extra requirements one-by-one. We exploit the fact that some cantons never imposed any further restriction then those required at a Federal level, while others mixed stricter face-mask requirements with further containment measures. In particular, we consider contact tracing policies (e.g. requirement of collecting people information in bars, restaurants, and other public spaces) and distancing policies (e.g. business closing, limiting the gathering of people, and schools closures). We construct three mutually-exclusive groups of policies (*Group*_*i*_) in which cantons are allocated according to the additional measures they adopted with respect to the Federal policy (Group 0). Namely, in Group 1 there are cantons enacting additional face-mask and tracing policies (but not distancing). In Group 2 there are cantons enacting additional face-mask and distancing policy (but not tracing). Finally, Group 3 includes those cantons that enforced all the three extra-measures. Formally, we estimate the following equation:

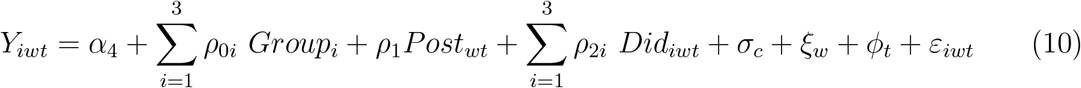

Where each *Group*_*i*_ is a dummy that takes value 1 according to whether the canton belongs to a specific policy group *i. Post*_*wt*_ is equal to 1 for the periods after the implementation of the policy. The parameters of interest are the *ρ*_2*i*_ for each of the three group. *ρ*_2*i*_ estimates the effect of the policy on group *i* with respect to group 0 (that is federal policy). Then, *Did*_*iwt*_ = *Group*_*i*_ *X Post*_*wt*_. Finally, *σ*_*c*_, *ξ*_*w*_ and *φ*_*t*_ are canton and time fixed-effects. In Table (11) we report the results of the estimation for the outcome at an aggregate age level.^5^ The policy-mix analysis confirms the overall small effect of the different policies. The heterogeneous achievements of the policies across genders are as well confirmed. Indeed, we find a marginally significant (*p* − *value <* 0.1) decrease in male mortality after the policy implementation of about 7%, irrespective of which group of policy is considered (−1.2 mask and tracing; -1.26 mask and distancing; -1.15 mask, tracing and distancing together). These latter results suggest that enacting additional distancing and tracing policies does not lead to better outcomes. The estimates on the two others group of policies are insignificant. In Table (11) we also report the results of the test run on the linear combination of the coefficients. The presumption here is that the effects are additively separable, admittedly a strong assumption, so that one can compute the separate effect of tracing by *Tracing* = *Did*(3) − *Did*(2) and similarly for *Mask* as noted in the table. We then test the effect of the singular NPI as a linear combination with the two others. Additional mask requirements decrease mortality for males (−1.31), very similar to our main estimate, but not for females. Tracing does not affect the outcome mean in any gender class. Finally, distancing policy has negligible and insignificant effects: +0.05 for male and -0.75 for females. These findings confirm that compulsory face-mask policy differently affects males and females. In addition, the policy-mix implemented doesn’t appear to achieve better outcomes than just wearing a face-mask in public places.

**Table 11:** Policy-mix analysis. Difference-in-Difference regression on share deaths.

| VARIABLES | (1)<br>Male | (2)<br>Male | (3)<br>Male | (4)<br>Female | (5)<br>Female | (6)<br>Female | (7)<br>Total | (8)<br>Total | (9)<br>Total |
| --- | --- | --- | --- | --- | --- | --- | --- | --- | --- |
| *Mask + Tracing*<sup>(a)</sup> | -1.46***<br>(0.42) | 0.20***<br>(0.01) | 0.20***<br>(0.01) | -2.80***<br>(0.50) | -0.79***<br>(0.01) | -0.79***<br>(0.01) | -2.13***<br>(0.46) | -0.30***<br>(0.00) | -0.30***<br>(0.00) |
| Did(1) | -1.20**<br>(0.48) | -1.20**<br>(0.48) | -1.20**<br>(0.48) | 1.17*<br>(0.63) | 1.17*<br>(0.63) | 1.17*<br>(0.63) | -0.02<br>(0.26) | -0.02<br>(0.26) | -0.02<br>(0.26) |
| *Mask + Distancing*<sup>(b)</sup> | 1.86***<br>(0.42) | 3.52***<br>(0.01) | 3.52***<br>(0.01) | 1.96***<br>(0.50) | 3.97***<br>(0.01) | 3.97***<br>(0.01) | 1.92***<br>(0.46) | 3.75***<br>(0.00) | 3.75***<br>(0.00) |
| Did(2) | -1.26**<br>(0.48) | -1.26**<br>(0.48) | -1.26**<br>(0.48) | -0.94<br>(0.63) | -0.94<br>(0.63) | -0.94<br>(0.63) | -1.12***<br>(0.26) | -1.12***<br>(0.26) | -1.12***<br>(0.26) |
| *Mask + Trac + Dist*<sup>(c)</sup> | 0.62<br>(0.94) | 0.54***<br>(0.01) | 0.54***<br>(0.01) | 0.78<br>(1.27) | 1.59***<br>(0.01) | 1.59***<br>(0.01) | 0.71<br>(1.09) | 1.07***<br>(0.01) | 1.07***<br>(0.01) |
| Did(3) | -1.15*<br>(0.67) | -1.15*<br>(0.67) | -1.15*<br>(0.67) | 0.42<br>(0.85) | 0.42<br>(0.85) | 0.42<br>(0.85) | -0.37<br>(0.49) | -0.37<br>(0.49) | -0.37<br>(0.49) |
| *Post*<sup>(d)</sup> | -0.67<br>(0.48) | -0.67<br>(0.48) | 0.09<br>(0.59) | -1.98***<br>(0.63) | -1.98***<br>(0.63) | -0.45<br>(0.62) | -1.32***<br>(0.26) | -1.32***<br>(0.26) | -0.16<br>(0.33) |
| Constant | 15.45***<br>(0.42) | 13.79***<br>(0.01) | 16.75***<br>(0.31) | 15.91***<br>(0.50) | 13.90***<br>(0.01) | 16.66***<br>(0.47) | 15.68***<br>(0.46) | 13.85***<br>(0.00) | 16.71***<br>(0.26) |
| Observations | 21,840 | 21,840 | 21,840 | 21,840 | 21,840 | 21,840 | 21,840 | 21,840 | 21,840 |
| R-squared | 0.01 | 0.08 | 0.12 | 0.01 | 0.12 | 0.19 | 0.02 | 0.17 | 0.26 |
| Year FE | NO | NO | YES | NO | NO | YES | NO | NO | YES |
| Canton FE | NO | YES | YES | NO | YES | YES | NO | YES | YES |
| Week FE | NO | NO | YES | NO | NO | YES | NO | NO | YES |
| *Mask*<sup>(e)</sup> | -1.31 | -1.31 | -1.31 | -0.19 | -0.19 | -0.19 | -0.77 | -0.77 | -0.77 |
| p-value | 0.06 | 0.06 | 0.82 | 0.02 | 0.83 | 0.20 | 0.40 | 0.40 | 0.40 |
| *Tracing*<sup>(f)</sup> | 0.11 | 0.11 | 0.11 | 1.36 | 1.36 | 1.36 | 0.75 | 0.75 | 0.75 |
| p-value | 0.82 | 0.82 | 0.06 | 0.20 | 0.02 | 0.83 | 0.08 | 0.08 | 0.08 |
| *Distancing*<sup>(g)</sup> | 0.05 | 0.05 | 0.05 | -0.75 | -0.75 | -0.75 | -0.35 | -0.35 | -0.35 |
| p-value | 0.92 | 0.92 | 0.92 | 0.83 | 0.20 | 0.02 | 0.13 | 0.13 | 0.13 |

Note: Observations: 21,840. Results of regression in equation 10 Dependent variable defined as Equation (1). (a) Cantons in treatment group 1: FR. Did(1) = Group1 X Post; (b) Cantons in treatment group 2: NA, Did(2) = Group2 X Post; (c) Cantons in treatment group 3: BS GE JU SO VD VS ZH, Did(3) = Group3 X Post. (d) *Post* is equal to 1 for all cantons after July, 7. S.E. clustered at a canton level (\*\*\* p<0.01, \*\* p<0.05, \* p<0.1). (e) Point estimate of the Test $H_0 : (Did3 - Did2) - Did1 \times (-1) = 0$ . (f) Point estimate of the Test $H_0 : (Did3 - Did2) = 0$ . (g) Point estimate of the Test $H_0 : (Did3 - Did2) = 0$ . P-values in (e)-(g) are the significance levels at which $H_0$ is rejected.

## 7 Conclusion

In this paper we investigate the impact of commonly implemented NPIs in the fight against COVID-19. We use a number of research designs to assuage different types of concerns regarding the plausibility of our estimated effects. Overall, our results suggest that extending mandatory face mask wearing to public indoor places (e.g. supermarkets, stations, airports, etc.) has a small and marginally significant effect on males only. We find a 5 − 10% reduction in male mortality using a diff-in-diff approach, however the results are not particularly robust. For example, in the event study we do not find any significant effect for either gender overall or by age cohort. We identify two likely source of potential heterogeneity. First, we account for the demographic structure of the population in terms of age and gender. Second, we investigate the effect of different policy-mix implementation. Overall no additional benefits appear from imposing additional measures (e.g. tracing). However, our findings do not suggest that face masks are ineffective, we can only test for the additional effect of imposing compulsory mask wearing in public places on top of public transportation. There are some caveats in the present work. First, even with administrative data deaths might be recorded with some delay (we have however checked for data updates). Second, it is plausible that our estimates are lower bound estimates of the effects if control cantons behave as *de facto* treated ones, i.e. if people consistently wear masks irrespective of cantonal rules. Future research should investigate in detail other aspects of the pandemic due to overwhelmed health facilities, such as hospital and medical treatment access and other specific causes of death different than Covid-19.

## Data Availability

Data all fully publicly available and we provide all the guidelines to download those data from the Swiss National Statistical Office.

## APPENDIX A

### A.1 Summary Statistics by canton

**Table A1:** Share Deaths^*a*^ per canton and sex

| Canton | Male | Female | Total | Canton | Male | Female | Total |
| --- | --- | --- | --- | --- | --- | --- | --- |
| AG | 13.782<br>(2.409) | 13.873<br>(2.575) | 13.827<br>(1.997) | OW | 14.242<br>(9.124) | 14.131<br>(9.167) | 14.189<br>(6.602) |
| AI | 16.211<br>(14.372) | 16.747<br>(15.444) | 16.474<br>(10.770) | SG | 14.843<br>(2.835) | 15.606<br>(3.247) | 15.227<br>(2.445) |
| AR | 16.656<br>(8.319) | 18.024<br>(8.547) | 17.338<br>(6.189) | SH | 17.529<br>(6.879) | 18.866<br>(6.977) | 18.212<br>(5.065) |
| BE | 17.490<br>(2.430) | 18.090<br>(2.903) | 17.797<br>(2.285) | SO | 16.484<br>(3.891) | 17.232<br>(4.293) | 16.862<br>(3.121) |
| BL | 15.690<br>(3.522) | 15.314<br>(3.891) | 15.498<br>(2.835) | SZ | 13.735<br>(4.653) | 13.583<br>(4.818) | 13.661<br>(3.374) |
| BS | 20.011<br>(5.253) | 23.441<br>(5.802) | 21.803<br>(4.312) | TG | 14.442<br>(3.778) | 14.735<br>(3.939) | 14.588<br>(2.968) |
| FR | 13.968<br>(3.692) | 13.100<br>(3.740) | 13.532<br>(2.874) | TI | 16.721<br>(4.114) | 17.244<br>(4.184) | 16.992<br>(3.457) |
| GE | 13.105<br>(2.741) | 13.706<br>(2.836) | 13.417<br>(2.208) | UR | 16.995<br>(10.596) | 17.150<br>(10.458) | 17.070<br>(7.556) |
| GL | 17.982<br>(9.678) | 19.250<br>(10.555) | 18.617<br>(7.205) | VD | 14.616<br>(2.798) | 14.874<br>(2.888) | 14.748<br>(2.412) |
| GR | 16.545<br>(4.499) | 16.763<br>(4.634) | 16.654<br>(3.361) | VS | 15.979<br>(3.650) | 14.594<br>(3.733) | 15.277<br>(2.879) |
| JU | 17.798<br>(7.377) | 17.316<br>(7.037) | 17.553<br>(5.220) | ZG | 11.710<br>(4.560) | 12.466<br>(4.933) | 12.085<br>(3.462) |
| LU | 14.463<br>(3.075) | 14.760<br>(3.478) | 14.613<br>(2.571) | ZH | 14.308<br>(2.027) | 15.468<br>(2.400) | 14.896<br>(1.930) |
| NE | 17.288<br>(4.822) | 17.828<br>(4.905) | 17.564<br>(3.707) | Total | 15.619<br>(6.400) | 16.058<br>(6.840) | 15.843<br>(5.071) |
| NW | 13.507<br>(7.886) | 13.346<br>(8.624) | 13.427<br>(6.053) |  |  |  |  |

Note: Observations 21840. Summary statistics of *ShareDeaths* for each canton. Each row contains the mean of the variable at an aggregated age-class level (0 to 90+ years old) for a specific sex in the first 40 weeks of the period January 2000 - October 2020. Standard deviation in parenthesis. (a) *Share Deaths* is the ratio between total deaths in year $t$ and population in $t - 1$ . Variables are expressed in 100.000 ratio.

### A.2 Event study by sex and age-class

**Figure A1:**
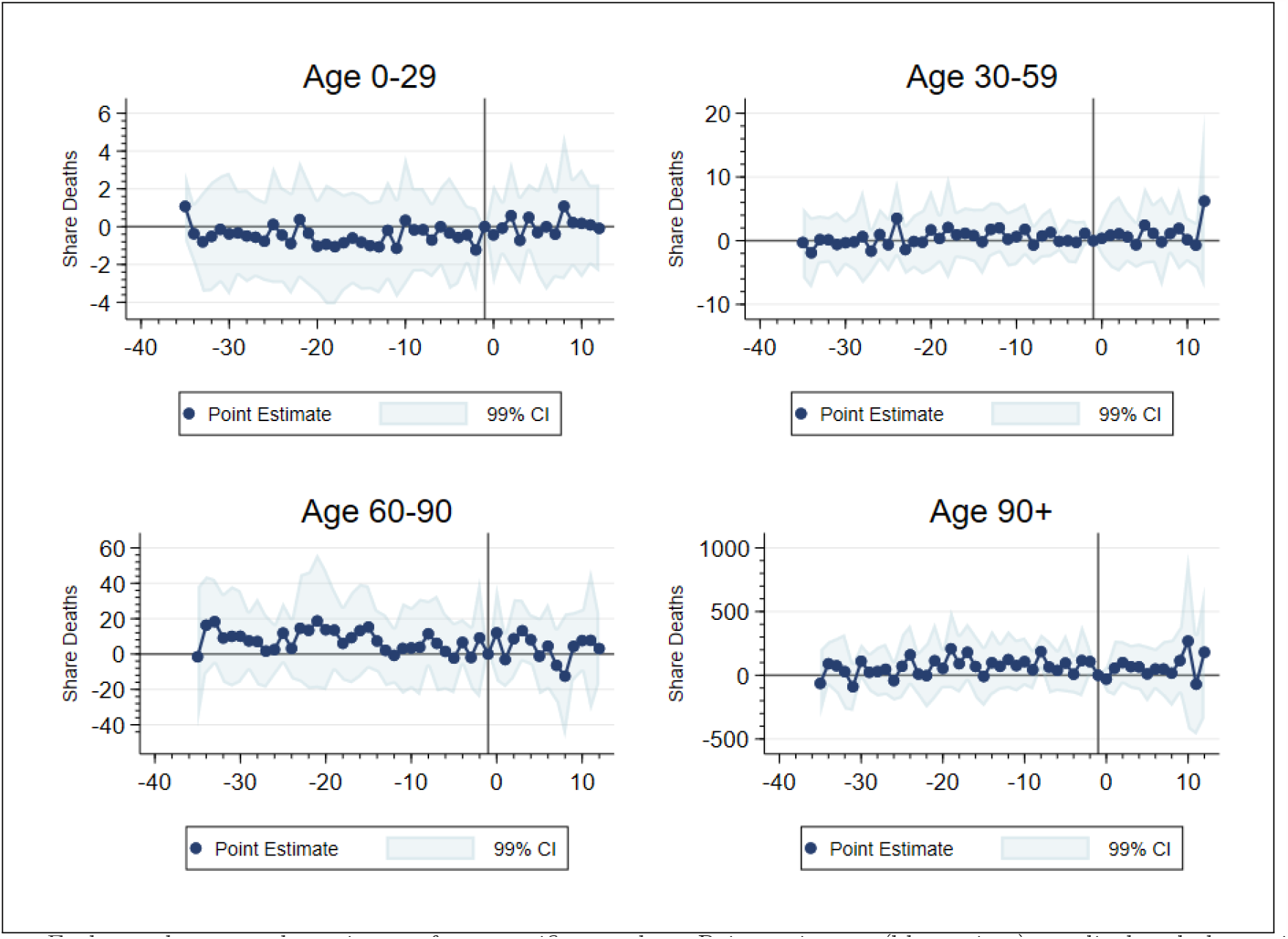
Share Deaths of Males by Age-Class Notes: Each panel reports the estimates for a specific age-class. Point estimates (blue points) are displayed along with their 99% confidence intervals (light blue area). Baseline period: 1 week prior to the adoption of the face masks policy in each adopting canton, corresponding to the solid vertical line in the plot.

**Figure A2:**
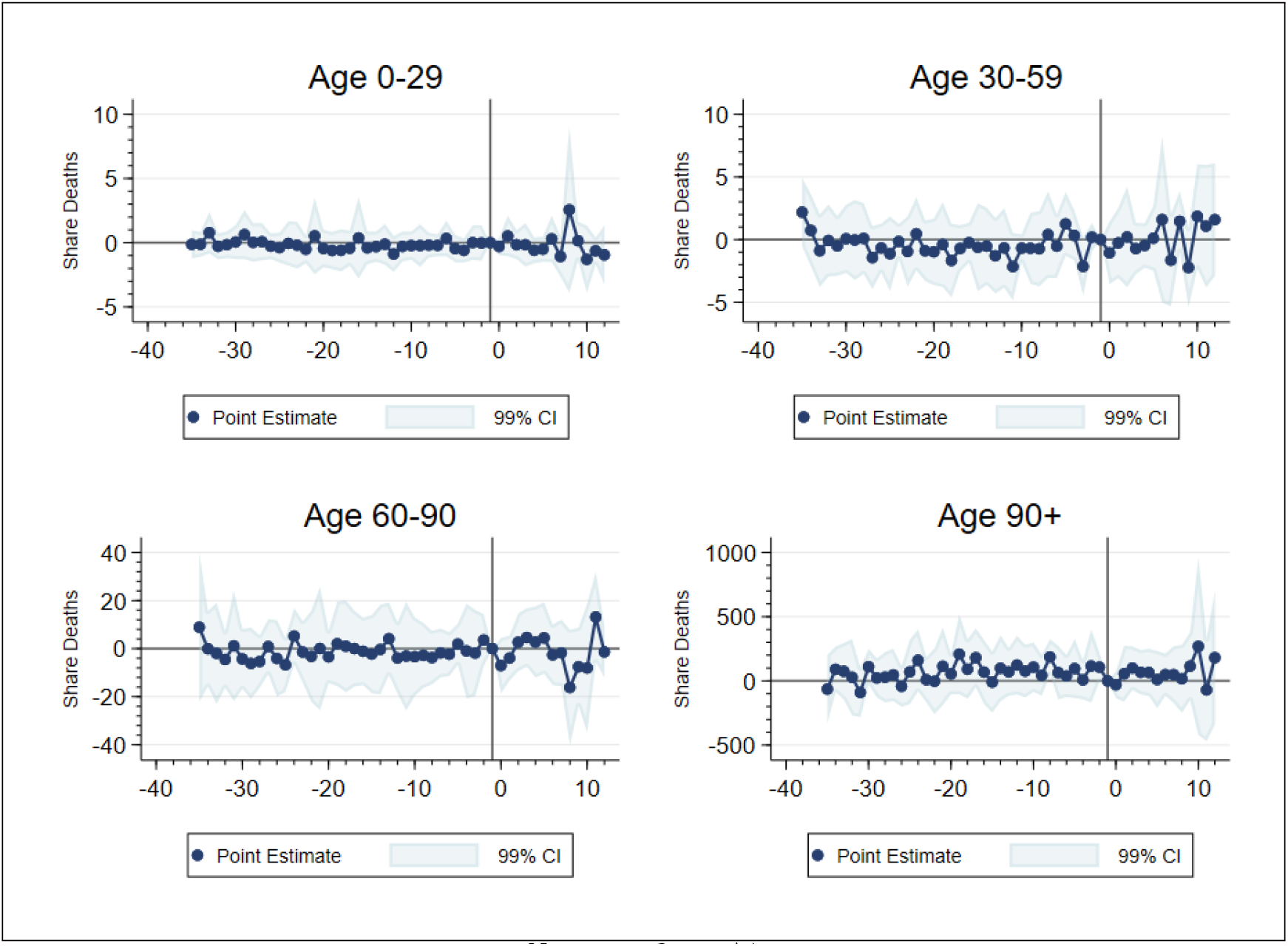
Share Deaths of Females by Age-Class Notes: see figure A1

**Figure A3:**
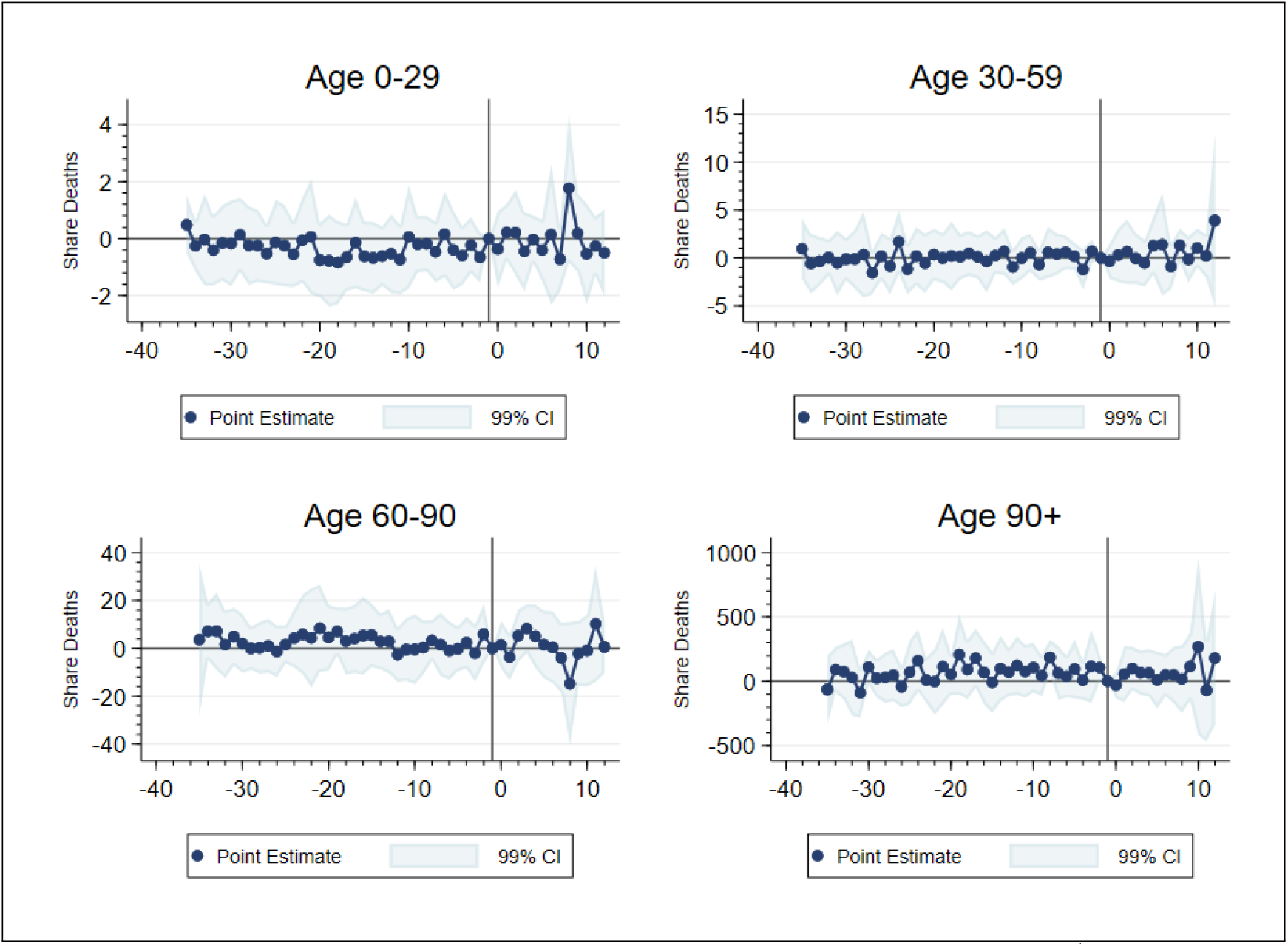
Share Deaths of Total Population by Age-Class Notes: Total population is aggregated male and female. For more details see A1

### A.3 Dynamic Betas by sex and age-class

**Figure A4:**
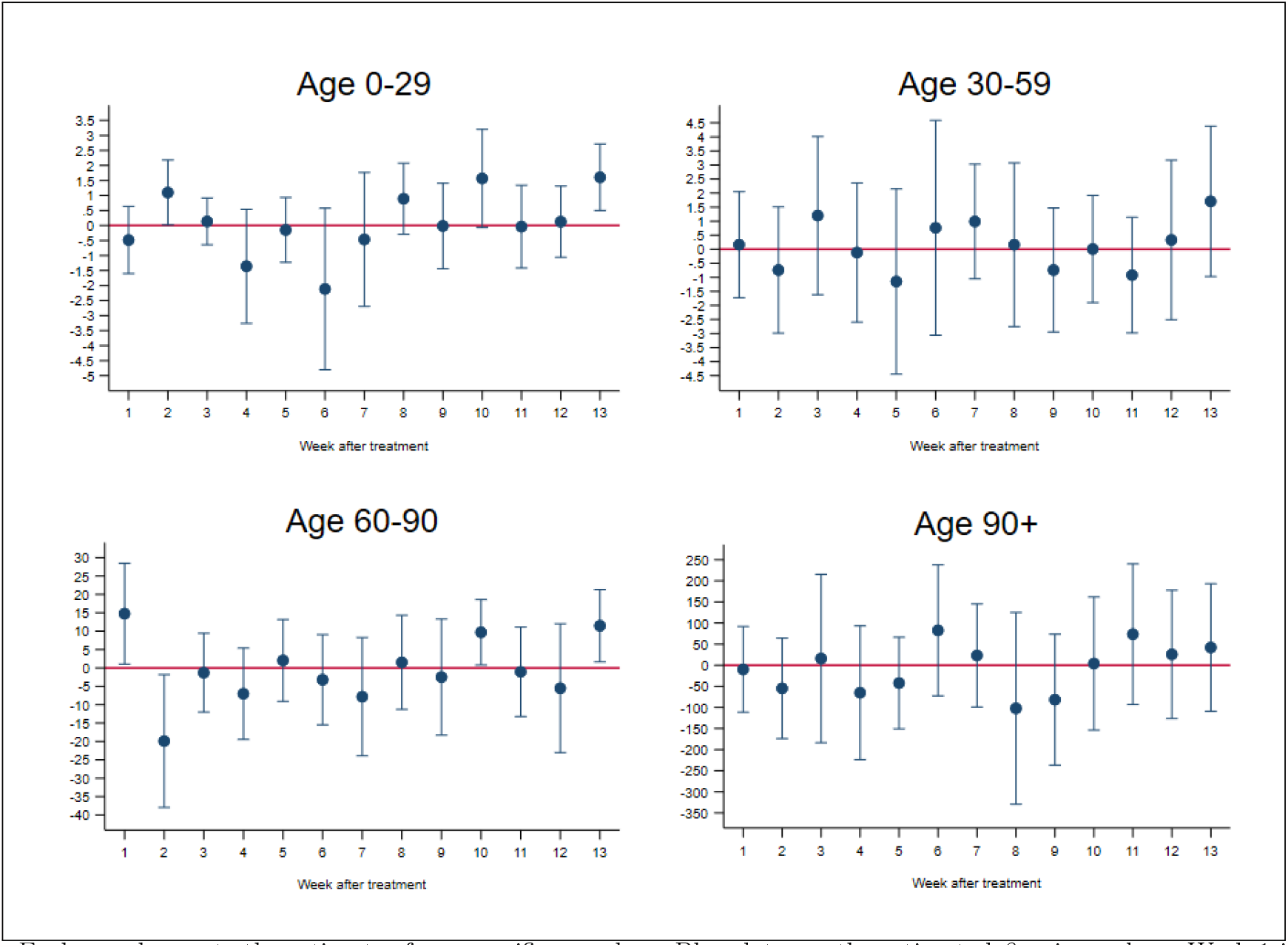
Difference-in-Difference Regression on Male Deaths with Dynamic Beta, by Age Class Note: Each panel reports the estimates for a specific age-class. Blue dots are the estimated *β*_2*w*_ in week *w*. Week 1 is the first week after the treatment, until the 13^*th*^ week after the treatment. Outcome defined as equation (2) expressed in 100.000. Each bar represents the clustered SE of the point estimates (*** p*<*0.01, ** p*<*0.05, * p*<*0.1).

**Figure A5:**
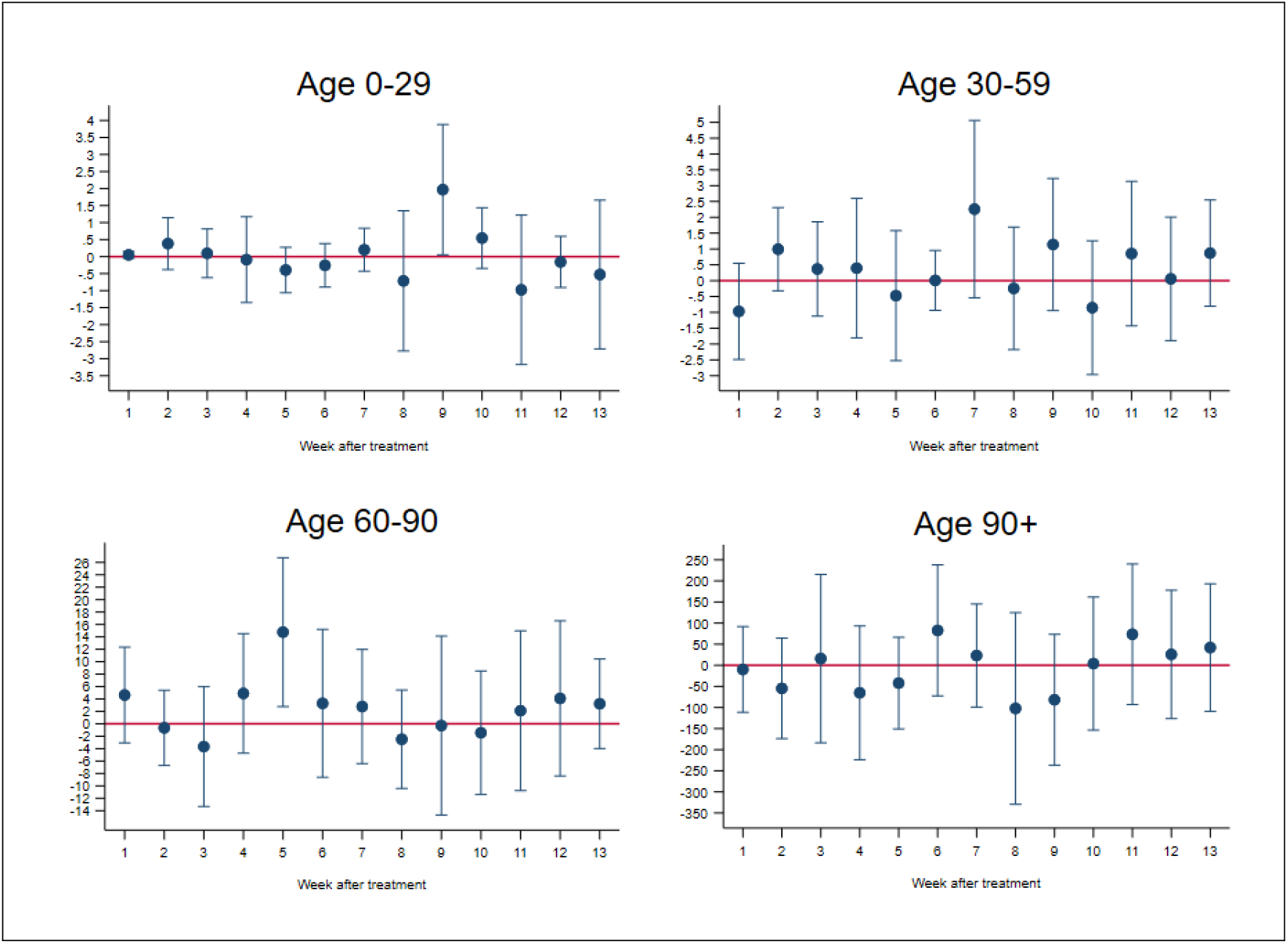
Difference-in-Difference Regression on Female Deaths with Dynamic Beta, by Age Class Note: see figure A4.

**Figure A6:**
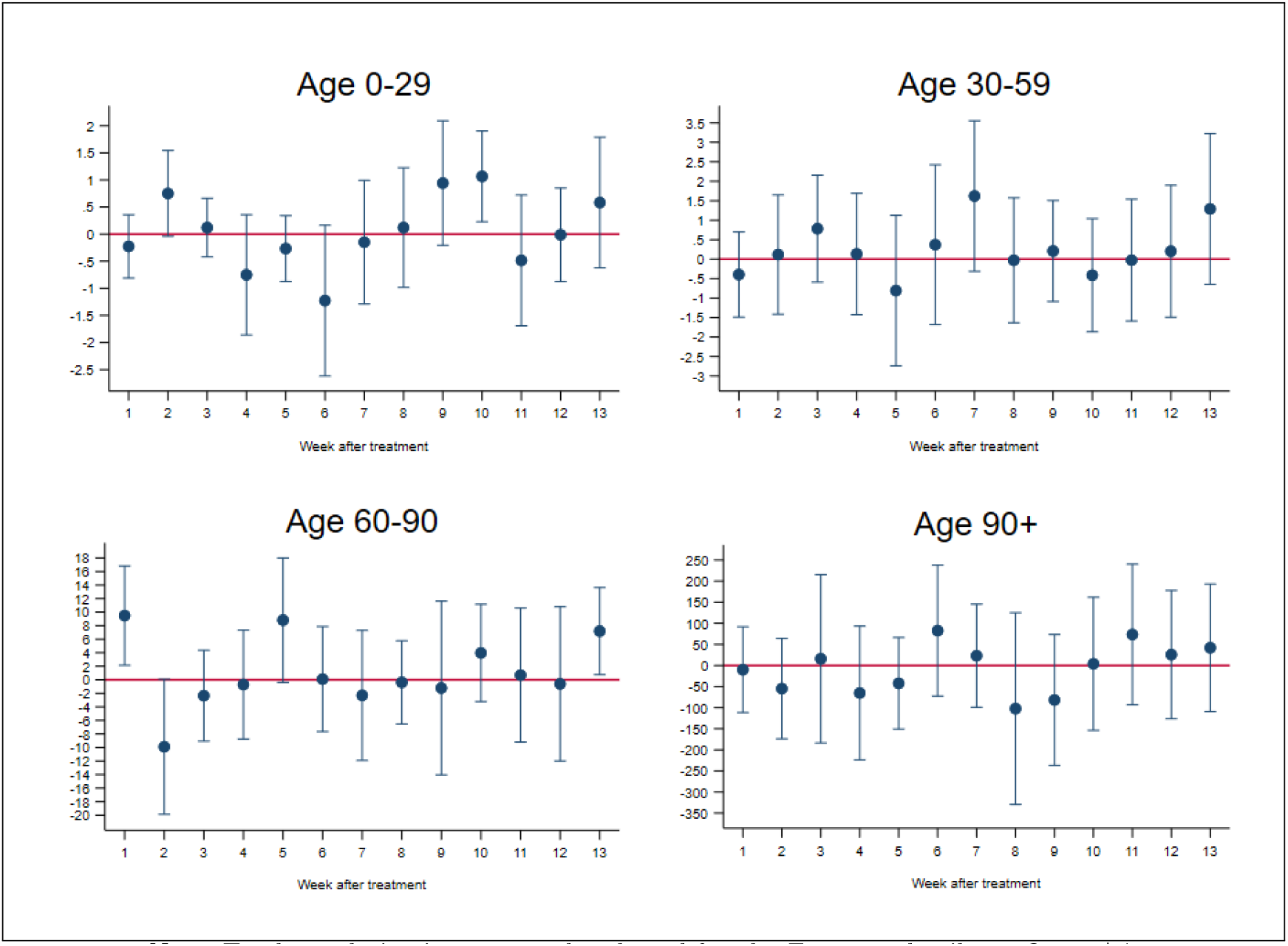
Difference-in-Difference Regression on Total Deaths with Dynamic Beta, by Age Class Note: Total population is aggregated male and female. For more details see figure A4.

### A.5 Pooled regression on sex

**Table A2:**
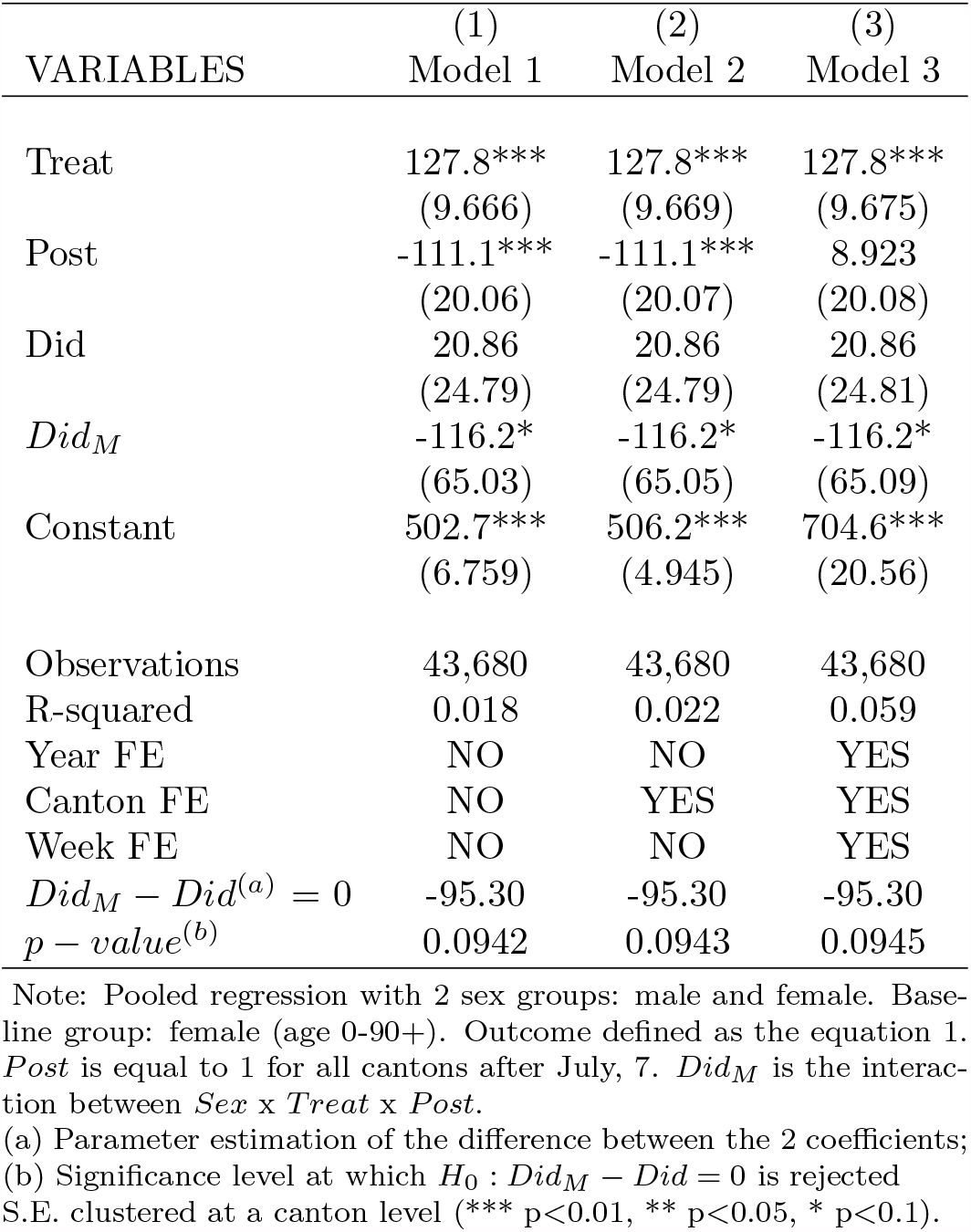
Pooled Regression with sex

### A.6 Difference-in-Difference with no pre-trend constrains

**Table A3:**
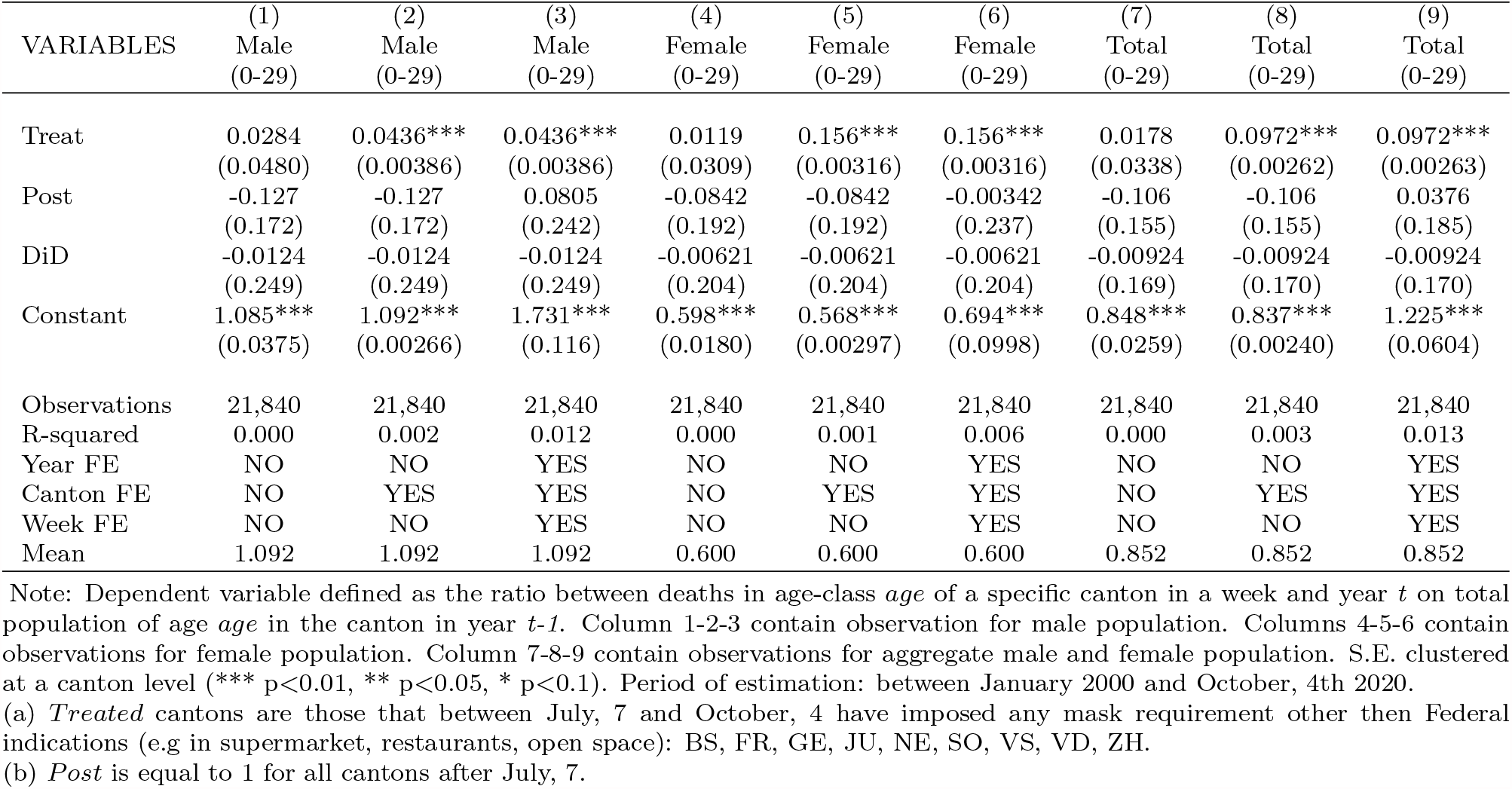
Difference-in-Difference share deaths with no pre-trend constrains: age 0-29

| VARIABLES | (1)<br>Male<br>(0-29) | (2)<br>Male<br>(0-29) | (3)<br>Male<br>(0-29) | (4)<br>Female<br>(0-29) | (5)<br>Female<br>(0-29) | (6)<br>Female<br>(0-29) | (7)<br>Total<br>(0-29) | (8)<br>Total<br>(0-29) | (9)<br>Total<br>(0-29) |
| --- | --- | --- | --- | --- | --- | --- | --- | --- | --- |
| Treat | 0.0284<br>(0.0480) | 0.0436***<br>(0.00386) | 0.0436***<br>(0.00386) | 0.0119<br>(0.0309) | 0.156***<br>(0.00316) | 0.156***<br>(0.00316) | 0.0178<br>(0.0338) | 0.0972***<br>(0.00262) | 0.0972***<br>(0.00263) |
| Post | -0.127<br>(0.172) | -0.127<br>(0.172) | 0.0805<br>(0.242) | -0.0842<br>(0.192) | -0.0842<br>(0.192) | -0.00342<br>(0.237) | -0.106<br>(0.155) | -0.106<br>(0.155) | 0.0376<br>(0.185) |
| DiD | -0.0124<br>(0.249) | -0.0124<br>(0.249) | -0.0124<br>(0.249) | -0.00621<br>(0.204) | -0.00621<br>(0.204) | -0.00621<br>(0.204) | -0.00924<br>(0.169) | -0.00924<br>(0.170) | -0.00924<br>(0.170) |
| Constant | 1.085***<br>(0.0375) | 1.092***<br>(0.00266) | 1.731***<br>(0.116) | 0.598***<br>(0.0180) | 0.568***<br>(0.00297) | 0.694***<br>(0.0998) | 0.848***<br>(0.0259) | 0.837***<br>(0.00240) | 1.225***<br>(0.0604) |
| Observations | 21,840 | 21,840 | 21,840 | 21,840 | 21,840 | 21,840 | 21,840 | 21,840 | 21,840 |
| R-squared | 0.000 | 0.002 | 0.012 | 0.000 | 0.001 | 0.006 | 0.000 | 0.003 | 0.013 |
| Year FE | NO | NO | YES | NO | NO | YES | NO | NO | YES |
| Canton FE | NO | YES | YES | NO | YES | YES | NO | YES | YES |
| Week FE | NO | NO | YES | NO | NO | YES | NO | NO | YES |
| Mean | 1.092 | 1.092 | 1.092 | 0.600 | 0.600 | 0.600 | 0.852 | 0.852 | 0.852 |

Note: Dependent variable defined as the ratio between deaths in age-class *age* of a specific canton in a week and year *t* on total population of age *age* in the canton in year *t-1*. Column 1-2-3 contain observation for male population. Columns 4-5-6 contain observations for female population. Column 7-8-9 contain observations for aggregate male and female population. S.E. clustered at a canton level (\*\*\*) $p < 0.01$ , \*\* $p < 0.05$ , \* $p < 0.1$ ). Period of estimation: between January 2000 and October, 4th 2020.
(a) *Treated* cantons are those that between July, 7 and October, 4 have imposed any mask requirement other then Federal indications (e.g in supermarket, restaurants, open space): BS, FR, GE, JU, NE, SO, VS, VD, ZH.
(b) *Post* is equal to 1 for all cantons after July, 7.

**Table A4:**
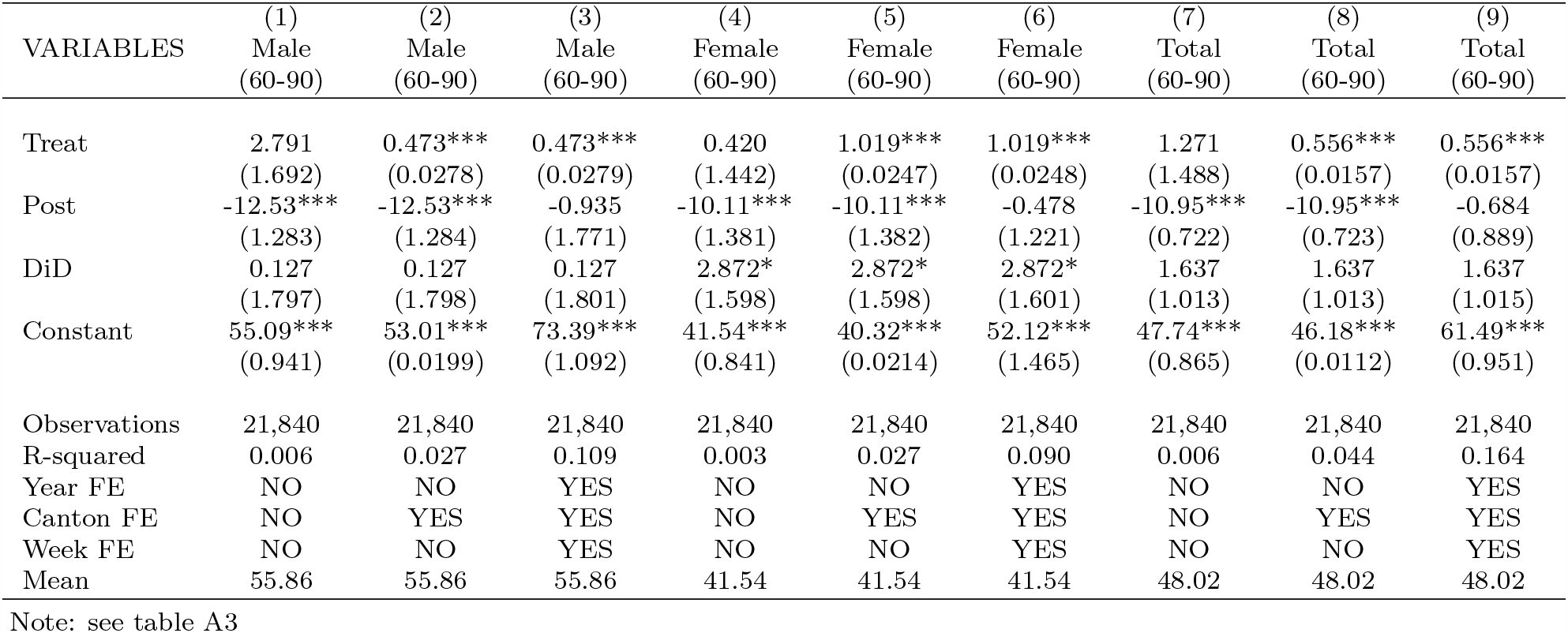
Difference-in-Difference share deaths with no pre-trend constrains: age 60-90

**Table A5:** Difference-in-Difference share deaths with no pre-trend constrains: age 90+

| VARIABLES | (1)<br>Male<br>(90+) | (2)<br>Male<br>(90+) | (3)<br>Male<br>(90+) | (4)<br>Female<br>(90+) | (5)<br>Female<br>(90+) | (6)<br>Female<br>(90+) | (7)<br>Total<br>(90+) | (8)<br>Total<br>(90+) | (9)<br>Total<br>(90+) |
| --- | --- | --- | --- | --- | --- | --- | --- | --- | --- |
| Treat | -22.85<br>(14.18) | -20.14***<br>(0.843) | -20.14***<br>(0.845) | -21.82*<br>(11.17) | -25.54***<br>(0.369) | -25.54***<br>(0.369) | -20.51*<br>(10.74) | -25.07***<br>(0.309) | -25.07***<br>(0.310) |
| Post | -33.67<br>(52.54) | -33.67<br>(52.57) | 92.51<br>(59.54) | -100.1***<br>(19.01) | -100.1***<br>(19.02) | -9.167<br>(21.08) | -77.47***<br>(15.59) | -77.47***<br>(15.60) | 19.99<br>(18.65) |
| DiD | -95.54*<br>(54.47) | -95.54*<br>(54.50) | -95.54*<br>(54.57) | 17.65<br>(23.82) | 17.65<br>(23.83) | 17.65<br>(23.87) | -14.96<br>(19.97) | -14.96<br>(19.98) | -14.96<br>(20.01) |
| Constant | 570.6***<br>(10.94) | 565.8***<br>(0.814) | 758.5***<br>(40.02) | 458.4***<br>(6.394) | 473.6***<br>(0.294) | 642.7***<br>(22.45) | 484.0***<br>(6.592) | 498.6***<br>(0.241) | 674.3***<br>(16.84) |
| Observations | 21,840 | 21,840 | 21,840 | 21,840 | 21,840 | 21,840 | 21,840 | 21,840 | 21,840 |
| R-squared | 0.001 | 0.005 | 0.034 | 0.002 | 0.009 | 0.070 | 0.002 | 0.011 | 0.095 |
| Year FE | NO | NO | YES | NO | NO | YES | NO | NO | YES |
| Canton FE | NO | YES | YES | NO | YES | YES | NO | YES | YES |
| Week FE | NO | NO | YES | NO | NO | YES | NO | NO | YES |
| Mean | 561.7 | 561.7 | 561.7 | 449.3 | 449.3 | 449.3 | 475.6 | 475.6 | 475.6 |

Note: see table A3

### A.7 Staggered Difference-in-Difference

We estimate the following equation:

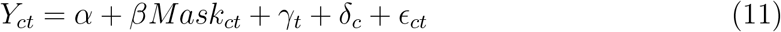

**Table A6:**
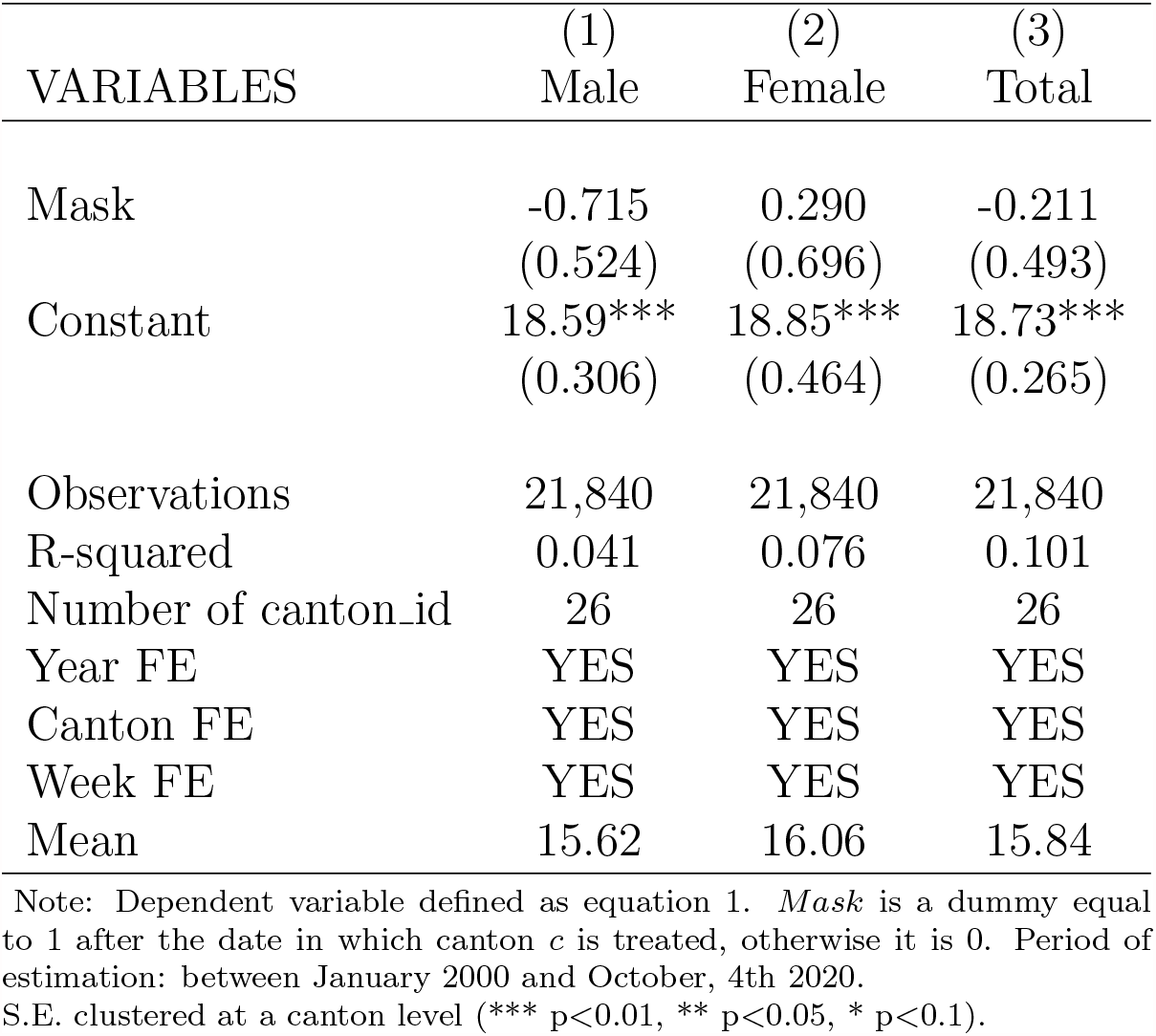
Staggered Difference-in-Difference

| VARIABLES | (1)<br>Male | (2)<br>Female | (3)<br>Total |
| --- | --- | --- | --- |
| Mask | -0.715<br>(0.524) | 0.290<br>(0.696) | -0.211<br>(0.493) |
| Constant | 18.59***<br>(0.306) | 18.85***<br>(0.464) | 18.73***<br>(0.265) |
| Observations | 21,840 | 21,840 | 21,840 |
| R-squared | 0.041 | 0.076 | 0.101 |
| Number of canton_id | 26 | 26 | 26 |
| Year FE | YES | YES | YES |
| Canton FE | YES | YES | YES |
| Week FE | YES | YES | YES |
| Mean | 15.62 | 16.06 | 15.84 |

Note: Dependent variable defined as equation 1. *Mask* is a dummy equal to 1 after the date in which canton *c* is treated, otherwise it is 0. Period of estimation: between January 2000 and October, 4th 2020. S.E. clustered at a canton level (\*\*\*) $p < 0.01$ , \*\* $p < 0.05$ , \* $p < 0.1$ .

**Table A7:** Staggered Difference-in-Difference: age 0-29

| VARIABLES | Male (0-29) | Female (0-29) | Total (0-29) |
| --- | --- | --- | --- |
| mask | 0.203<br>(0.233) | 0.140<br>(0.117) | 0.170<br>(0.103) |
| Constant | 1.108***<br>(0.127) | 0.567***<br>(0.104) | 0.844***<br>(0.0554) |
| Observations | 13,520 | 13,520 | 13,520 |
| R-squared | 0.005 | 0.004 | 0.004 |
| Number of canton_id | 26 | 26 | 26 |
| Year FE | YES | YES | YES |
| Canton FE | YES | YES | YES |
| Week FE | YES | YES | YES |
| Mean | 0.952 | 0.541 | 0.752 |

Note: Dependent variable defined as equation 2. For more details see A6

**Table A8:** Staggered Difference-in-Difference: age 30-59

| VARIABLES | Male (30-59) | Female (30-59) | Total (30-59) |
| --- | --- | --- | --- |
| mask | -0.0841<br>(0.324) | 0.296<br>(0.358) | 0.108<br>(0.313) |
| Constant | 5.423***<br>(0.300) | 2.737***<br>(0.208) | 4.103***<br>(0.157) |
| Observations | 21,840 | 21,840 | 21,840 |
| R-squared | 0.017 | 0.009 | 0.023 |
| Number of canton_id | 26 | 26 | 26 |
| Year FE | YES | YES | YES |
| Canton FE | YES | YES | YES |
| Week FE | YES | YES | YES |
| Mean | 3.905 | 2.307 | 3.115 |

Note: Dependent variable defined as equation 2. For more details see A6

**Table A9:**
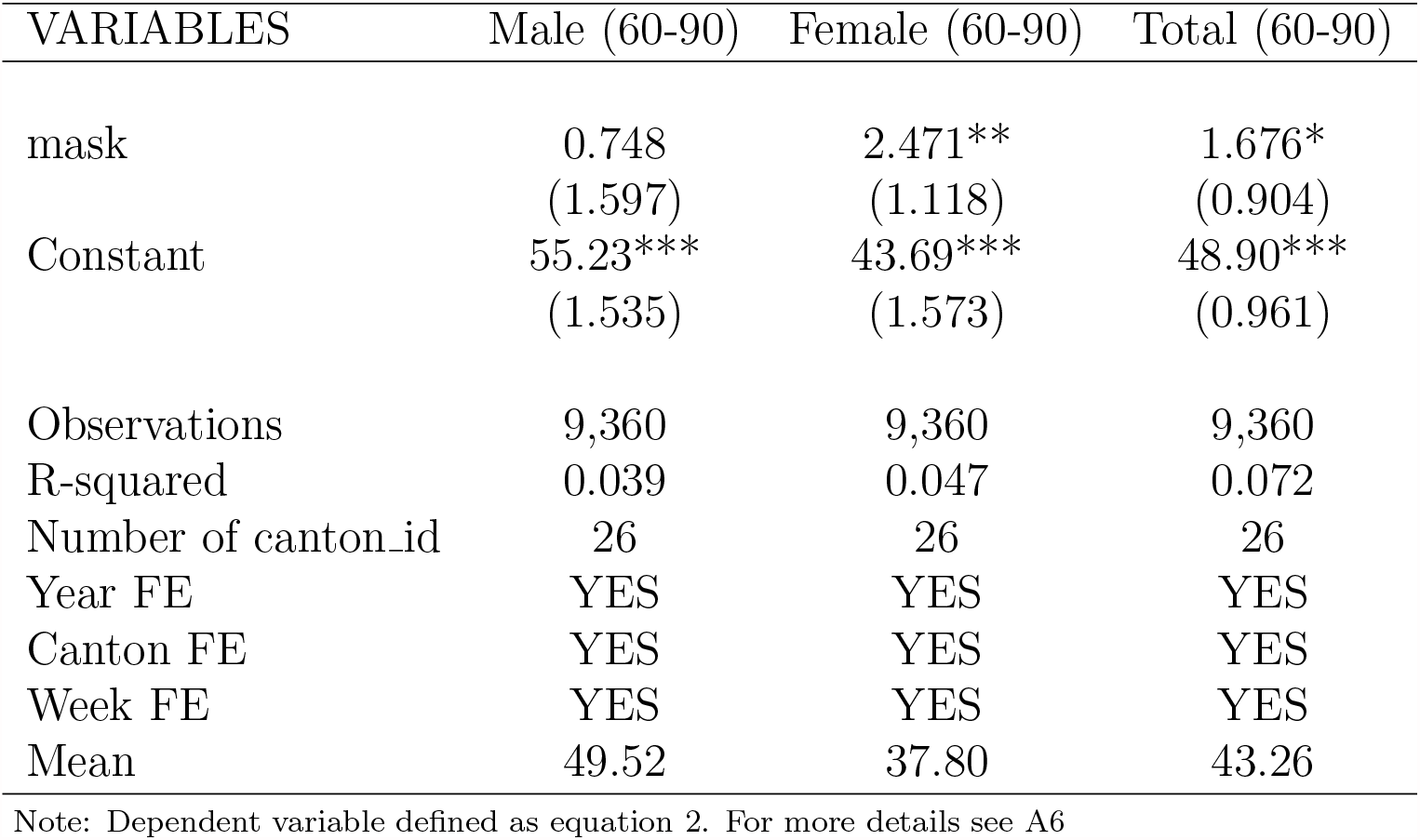
Staggered Difference-in-Difference: age 60-90

**Table A10:** Staggered Difference-in-Difference: age 90+

| VARIABLES | Male 90+ | Female 90+ | Total 90+ |
| --- | --- | --- | --- |
| mask | -47.09<br>(37.48) | 15.74<br>(17.45) | -0.700<br>(16.95) |
| Constant | 669.0***<br>(34.01) | 534.1***<br>(14.72) | 566.0***<br>(14.51) |
| Observations | 13,520 | 13,520 | 13,520 |
| R-squared | 0.035 | 0.066 | 0.094 |
| Number of canton_id | 26 | 26 | 26 |
| Year FE | YES | YES | YES |
| Canton FE | YES | YES | YES |
| Week FE | YES | YES | YES |
| Mean | 549.2 | 443 | 468.6 |

Note: Dependent variable defined as equation 2. For more details see A6

### A.8 Policy mix tables per age-class

**Table A11:** Policy Mix regression: age 0-29

| VARIABLES | (1)<br>Male<br>(0-29) | (2)<br>Male<br>(0-29) | (3)<br>Male<br>(0-29) | (4)<br>Female<br>(0-29) | (5)<br>Female<br>(0-29) | (6)<br>Female<br>(0-29) | (7)<br>Total<br>(0-29) | (8)<br>Total<br>(0-29) | (9)<br>Total<br>(0-29) |
| --- | --- | --- | --- | --- | --- | --- | --- | --- | --- |
| *Mask + Tracing*<sup>(a)</sup> | -0.06*<br>(0.03) | -0.01***<br>(0.00) | -0.01***<br>(0.00) | -0.05**<br>(0.02) | -0.05***<br>(0.00) | -0.05***<br>(0.00) | -0.06**<br>(0.03) | -0.03***<br>(0.00) | -0.03***<br>(0.00) |
| Diff(1) | 0.02<br>(0.17) | 0.02<br>(0.17) | 0.02<br>(0.17) | -0.05<br>(0.19) | -0.05<br>(0.20) | -0.05<br>(0.20) | -0.01<br>(0.15) | -0.01<br>(0.15) | -0.01<br>(0.15) |
| *Mask + Distancing*<sup>(b)</sup> | -0.06*<br>(0.03) | -0.01**<br>(0.00) | -0.01**<br>(0.00) | -0.00<br>(0.02) | -0.01<br>(0.00) | -0.01<br>(0.00) | -0.03<br>(0.03) | -0.01**<br>(0.00) | -0.01**<br>(0.00) |
| Diff(2) | -0.40**<br>(0.17) | -0.40**<br>(0.17) | -0.40**<br>(0.17) | 0.30<br>(0.19) | 0.30<br>(0.20) | 0.30<br>(0.20) | -0.06<br>(0.15) | -0.06<br>(0.15) | -0.06<br>(0.15) |
| *Mask + Trac + Dist*<sup>(c)</sup> | -0.04<br>(0.05) | -0.01<br>(0.01) | -0.01<br>(0.01) | 0.00<br>(0.04) | 0.08***<br>(0.01) | 0.08***<br>(0.01) | -0.02<br>(0.04) | 0.04***<br>(0.00) | 0.04***<br>(0.00) |
| Diff(3) | 0.13<br>(0.26) | 0.13<br>(0.26) | 0.13<br>(0.26) | -0.02<br>(0.21) | -0.02<br>(0.21) | -0.02<br>(0.21) | 0.06<br>(0.17) | 0.06<br>(0.17) | 0.06<br>(0.17) |
| Post | -0.01<br>(0.17) | -0.01<br>(0.17) | 0.02<br>(0.23) | -0.03<br>(0.19) | -0.03<br>(0.20) | 0.02<br>(0.24) | -0.02<br>(0.15) | -0.02<br>(0.15) | 0.02<br>(0.18) |
| Constant | 0.97***<br>(0.03) | 0.92***<br>(0.00) | 1.08***<br>(0.13) | 0.54***<br>(0.02) | 0.55***<br>(0.00) | 0.57***<br>(0.10) | 0.76***<br>(0.03) | 0.74***<br>(0.00) | 0.83***<br>(0.06) |
| Observations | 13,520 | 13,520 | 13,520 | 13,520 | 13,520 | 13,520 | 13,520 | 13,520 | 13,520 |
| R-squared | 0.00 | 0.00 | 0.01 | 0.00 | 0.00 | 0.01 | 0.00 | 0.00 | 0.01 |
| Year FE | NO | NO | YES | NO | NO | YES | NO | NO | YES |
| Canton FE | NO | YES | YES | NO | YES | YES | NO | YES | YES |
| Week FE | NO | NO | YES | NO | NO | YES | NO | NO | YES |
| *Mask*<sup>(e)</sup> | -0.51 | -0.51 | -0.51 | 0.27 | 0.27 | 0.27 | -0.13 | -0.13 | -0.13 |
| p-value | 0.06 | 0.01 | 0.06 | 0.79 | 0.00 | 0.79 | 0.46 | 0.47 | 0.37 |
| *Tracing*<sup>(f)</sup> | 0.53 | 0.53 | 0.53 | -0.32 | -0.32 | -0.32 | 0.11 | 0.11 | 0.11 |
| p-value | 0.01 | 0.06 | 0.02 | 0.00 | 0.79 | 0.21 | 0.13 | 0.13 | 0.47 |
| *Distancing*<sup>(g)</sup> | 0.11 | 0.11 | 0.11 | 0.02 | 0.02 | 0.02 | 0.07 | 0.07 | 0.07 |
| p-value | 0.58 | 0.58 | 0.58 | 0.21 | 0.21 | 0.00 | 0.37 | 0.37 | 0.13 |

Note: Observations: 21,840. Results of regression in equation 10. Dependent variable defined as Equation (1). (a) Cantons in treatment group 1: FR. Did(1) = Group1 X Post; (b) Cantons in treatment group 2: NA, Did(2) = Group2 X Post; (c) Cantons in treatment group 3: BS GE JU SO VD VS ZH, Did(3) = Group3 X Post. (d) Post is equal to 1 for all cantons after July, 7. S.E. clustered at a canton level (\*\*\*) p<0.01, \*\* p<0.05, \* p<0.1). (e) Point estimate of the Test $H_0: (Did3 - Did2 - Did1) X (-1) = 0$ . (f) Point estimate of the Test $H_0: (Did3 - Did2) = 0$ . (g) Point estimate of the Test $H_0: (Did3 - Did2) = 0$ . P-values in (e)-(g) are the significance levels at which $H_0$ is rejected.

**Table A12:** Policy Mix regression: age 30-59

| VARIABLES | (1)<br>Male<br>(30-59) | (2)<br>Male<br>(30-59) | (3)<br>Male<br>(30-59) | (4)<br>Female<br>(30-59) | (5)<br>Female<br>(30-59) | (6)<br>Female<br>(30-59) | (7)<br>Total<br>(30-59) | (8)<br>Total<br>(30-59) | (9)<br>Total<br>(30-59) |
| --- | --- | --- | --- | --- | --- | --- | --- | --- | --- |
| *Mask + Tracing<sup>(a)</sup>* | 0.42***<br>(0.10) | 0.53***<br>(0.00) | 0.53***<br>(0.00) | -0.04<br>(0.06) | -0.08***<br>(0.00) | -0.08***<br>(0.00) | 0.19**<br>(0.08) | 0.23***<br>(0.00) | 0.23***<br>(0.00) |
| Diff(1) | 0.61**<br>(0.27) | 0.61**<br>(0.27) | 0.61**<br>(0.27) | 0.26<br>(0.20) | 0.26<br>(0.20) | 0.26<br>(0.20) | 0.43**<br>(0.19) | 0.43**<br>(0.19) | 0.43**<br>(0.19) |
| *Mask + Distancing<sup>(b)</sup>* | 0.79***<br>(0.10) | 0.90***<br>(0.00) | 0.90***<br>(0.00) | 0.37***<br>(0.06) | 0.33***<br>(0.00) | 0.33***<br>(0.00) | 0.57***<br>(0.08) | 0.61***<br>(0.00) | 0.61***<br>(0.00) |
| Diff(2) | -0.25<br>(0.27) | -0.25<br>(0.27) | -0.25<br>(0.27) | -0.37*<br>(0.20) | -0.37*<br>(0.20) | -0.37*<br>(0.20) | -0.31<br>(0.19) | -0.31<br>(0.19) | -0.31<br>(0.19) |
| *Mask + Trac + Dist<sup>(c)</sup>* | 0.49**<br>(0.21) | -0.02***<br>(0.01) | -0.02***<br>(0.01) | 0.19<br>(0.12) | -0.09***<br>(0.01) | -0.09***<br>(0.01) | 0.33*<br>(0.16) | -0.05***<br>(0.00) | -0.05***<br>(0.00) |
| Diff(3) | 0.11<br>(0.33) | 0.11<br>(0.33) | 0.11<br>(0.33) | 0.45<br>(0.36) | 0.45<br>(0.36) | 0.45<br>(0.36) | 0.29<br>(0.30) | 0.29<br>(0.30) | 0.29<br>(0.30) |
| Post | -1.12***<br>(0.27) | -1.12***<br>(0.27) | -0.23<br>(0.28) | -0.79***<br>(0.20) | -0.79***<br>(0.20) | -0.29<br>(0.24) | -0.96***<br>(0.19) | -0.96***<br>(0.19) | -0.26<br>(0.23) |
| Constant | 3.74***<br>(0.10) | 3.63***<br>(0.00) | 5.13***<br>(0.30) | 2.25***<br>(0.06) | 2.29***<br>(0.00) | 2.71***<br>(0.21) | 3.01***<br>(0.08) | 2.97***<br>(0.00) | 3.94***<br>(0.16) |
| Observations | 21,840 | 21,840 | 21,840 | 21,840 | 21,840 | 21,840 | 21,840 | 21,840 | 21,840 |
| R-squared | 0.00 | 0.01 | 0.03 | 0.00 | 0.01 | 0.02 | 0.01 | 0.02 | 0.04 |
| Year FE | NO | NO | YES | NO | NO | YES | NO | NO | YES |
| Canton FE | NO | YES | YES | NO | YES | YES | NO | YES | YES |
| Week FE | NO | NO | YES | NO | NO | YES | NO | NO | YES |
| *Mask<sup>(e)</sup>* | 0.24 | 0.24 | 0.24 | -0.56 | -0.56 | -0.56 | -0.17 | -0.17 | -0.17 |
| p-value | 0.46 | 0.46 | 0.46 | 0.52 | 0.01 | 0.01 | 0.02 | 0.02 | 0.02 |
| *Tracing<sup>(f)</sup>* | 0.36 | 0.36 | 0.36 | 0.82 | 0.82 | 0.82 | 0.60 | 0.60 | 0.60 |
| p-value | 0.06 | 0.01 | 0.06 | 0.01 | 0.13 | 0.52 | 0.59 | 0.54 | 0.54 |
| *Distancing<sup>(g)</sup>* | -0.49 | -0.49 | -0.49 | 0.19 | 0.19 | 0.19 | -0.14 | -0.14 | -0.14 |
| p-value | 0.01 | 0.06 | 0.01 | 0.13 | 0.52 | 0.13 | 0.54 | 0.59 | 0.59 |

Note: see table A11

**Table A13:**
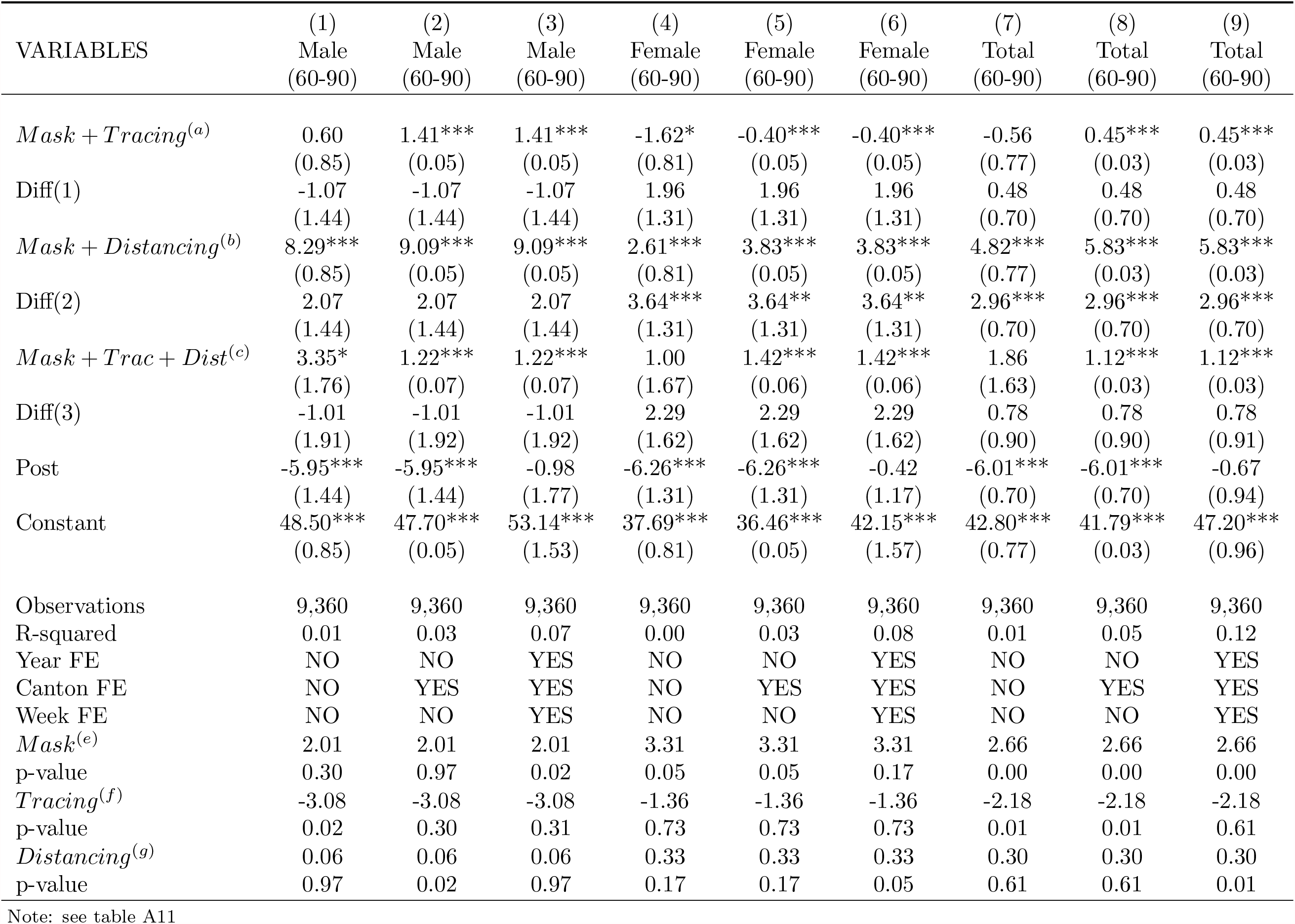
Policy Mix regression: age 60-90

**Table A14:** Policy Mix regression: age 90+

| VARIABLES | Male<br>(90+) | Male<br>(90+) | Male<br>(90+) | Female<br>(90+) | Female<br>(90+) | Female<br>(90+) | Total<br>(90+) | Total<br>(90+) | Total<br>(90+) |
| --- | --- | --- | --- | --- | --- | --- | --- | --- | --- |
| *Mask + Tracing*<sup>(a)</sup> | -11.75<br>(10.74) | -5.59***<br>(1.33) | -5.59***<br>(1.33) | -14.51**<br>(5.91) | -31.91***<br>(0.44) | -31.91***<br>(0.44) | -10.11<br>(6.25) | -26.22***<br>(0.40) | -26.22***<br>(0.40) |
| Diff(1) | -78.42<br>(53.17) | -78.42<br>(53.21) | -78.42<br>(53.31) | 112.47***<br>(17.49) | 112.47***<br>(17.51) | 112.47***<br>(17.54) | 57.07***<br>(15.93) | 57.07***<br>(15.95) | 57.07***<br>(15.98) |
| *Mask + Distancing*<sup>(b)</sup> | -12.57<br>(10.74) | -6.40***<br>(1.33) | -6.40***<br>(1.33) | -41.03***<br>(5.91) | -58.42***<br>(0.44) | -58.42***<br>(0.44) | -37.09***<br>(6.25) | -53.21***<br>(0.40) | -53.21***<br>(0.40) |
| Diff(2) | -171.55***<br>(53.17) | -171.55***<br>(53.21) | -171.55***<br>(53.31) | -54.23***<br>(17.49) | -54.23***<br>(17.51) | -54.23***<br>(17.54) | -85.78***<br>(15.93) | -85.78***<br>(15.95) | -85.78***<br>(15.98) |
| *Mask + Trac + Dist*<sup>(c)</sup> | -31.58**<br>(14.53) | -24.31***<br>(1.38) | -24.31***<br>(1.38) | -31.40**<br>(13.08) | -29.93***<br>(0.51) | -29.93***<br>(0.51) | -30.03**<br>(12.17) | -29.35***<br>(0.46) | -29.35***<br>(0.47) |
| Diff(3) | -81.45<br>(55.24) | -81.45<br>(55.29) | -81.45<br>(55.39) | 25.65<br>(20.46) | 25.65<br>(20.48) | 25.65<br>(20.52) | -4.72<br>(18.56) | -4.72<br>(18.57) | -4.72<br>(18.61) |
| Post | -23.00<br>(53.17) | -23.00<br>(53.21) | 90.22<br>(58.36) | -97.52***<br>(17.49) | -97.52***<br>(17.51) | -12.66<br>(20.80) | -73.86***<br>(15.93) | -73.86***<br>(15.95) | 17.32<br>(17.93) |
| Constant | 559.98***<br>(10.74) | 553.81***<br>(1.33) | 673.95***<br>(34.28) | 455.77***<br>(5.91) | 473.16***<br>(0.44) | 561.95***<br>(14.70) | 480.41***<br>(6.25) | 496.52***<br>(0.40) | 592.39***<br>(14.53) |
| Observations | 13,520 | 13,520 | 13,520 | 13,520 | 13,520 | 13,520 | 13,520 | 13,520 | 13,520 |
| R-squared | 0.00 | 0.01 | 0.04 | 0.00 | 0.01 | 0.07 | 0.00 | 0.01 | 0.10 |
| Year FE | NO | NO | YES | NO | NO | YES | NO | NO | YES |
| Canton FE | NO | YES | YES | NO | YES | YES | NO | YES | YES |
| Week FE | NO | NO | YES | NO | NO | YES | NO | NO | YES |
| *Mask*<sup>(e)</sup> | -168.51 | -168.51 | -168.51 | 32.58 | 32.58 | 32.58 | -23.99 | -23.99 | -23.99 |
| p-value | 0.01 | 0.84 | 0.00 | 0.12 | 0.12 | 0.12 | 0.21 | 0.21 | 0.21 |
| *Tracing*<sup>(f)</sup> | 90.10 | 90.10 | 90.10 | 79.89 | 79.89 | 79.89 | 81.06 | 81.06 | 81.06 |
| p-value | 0.00 | 0.00 | 0.01 | 0.00 | 0.00 | 0.00 | 0.00 | 0.00 | 0.00 |
| *Distancing*<sup>(g)</sup> | -3.03 | -3.03 | -3.03 | -86.81 | -86.81 | -86.81 | -61.79 | -61.79 | -61.79 |
| p-value | 0.84 | 0.01 | 0.84 | 0.00 | 0.00 | 0.00 | 0.00 | 0.00 | 0.00 |

Note: see table A11

## APPENDIX B

### B.1 Analysis on covid outcomes

Here we report the estimates of the Difference-in-Difference (equation (3)) on two covid deaths and cases. We both use the main identification strategy (table B2) and the dynamic *β* estimates (figures B3-B4). We narrow the period of analysis at year 2020 and add fixed effect accordingly. Figures (B2)-(B1) reports the parallel trends. For covid variables we do not have the demographic characteristic per canton of deaths. Thus, the outcome in this section is defined as follow:

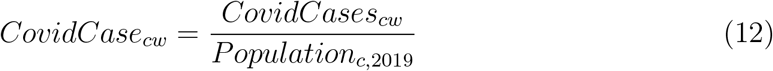

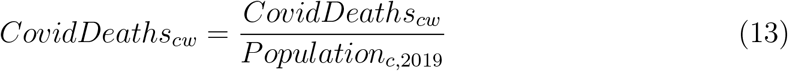

Where, *CovidCase*_*cw*_ and *CovidDeaths*_*cw*_ are, respectively, weekly case and the deaths in a specific canton in the first 40^*th*^ weeks of 2010. Both variables are weighted with the total population of the canton in the previous year (2019). Table (B1) reports the summary statistics for the two variables.

**Table B1:** Descriptive Statistics of Covid-19 variables

| Variable | Mean | Sd | Min | Max |
| --- | --- | --- | --- | --- |
| Aggregated Covid Deaths | 63.55 | 91.44 | 0 | 350 |
| Weekly Covid Deaths | 2.545 | 8.396 | 0 | 84 |
| Aggregated Covid Case | 1304.5 | 1789.6 | 3 | 10885 |
| Weekly Covid Case | 70.73 | 163.1 | 0 | 1418 |
| Share covid case | 17.40 | 29.45 | 0 | 272.5 |
| Share covid deaths | 0.714 | 2.111 | 0 | 23.90 |
| Observations<sup>(a)</sup> | 754 |  |  |  |

Note: (a) Observations in the first 40 weeks of year 2020. Summary statistics of covid-related variables for whole Switzerland. Each row contains the mean of the variable at an aggregate age-class level. *Aggregated Covid Deaths* and *Aggregated Covid Case* are the cumulative number in each week. *Share Deaths* and *Share Case* are the ratio between covid deaths/cases in $y$ and population in the same year ( $y$ ), per 100.000.

With linear diff-in-diff we do not find any statistical significant results. However, the estimated coefficient for covid deaths share is negative (between -0.68 and -0.62), while it is positive for covid new cases (between +1.5 and +2). The ineffectiveness of the policy on covid-outcomes is also confirmed with the the dynamic *β* specification. Indeed, covid cases undergo a constant increase between the 1st and the 10th week after the implementation of the policy, decreasing in week 11 and 12 and increasing again in week 13. On the other hand, covid deaths remain more or less stable, but the estimate are quite imprecise due to high standard errors around the estimated points.

**Figure B1:**
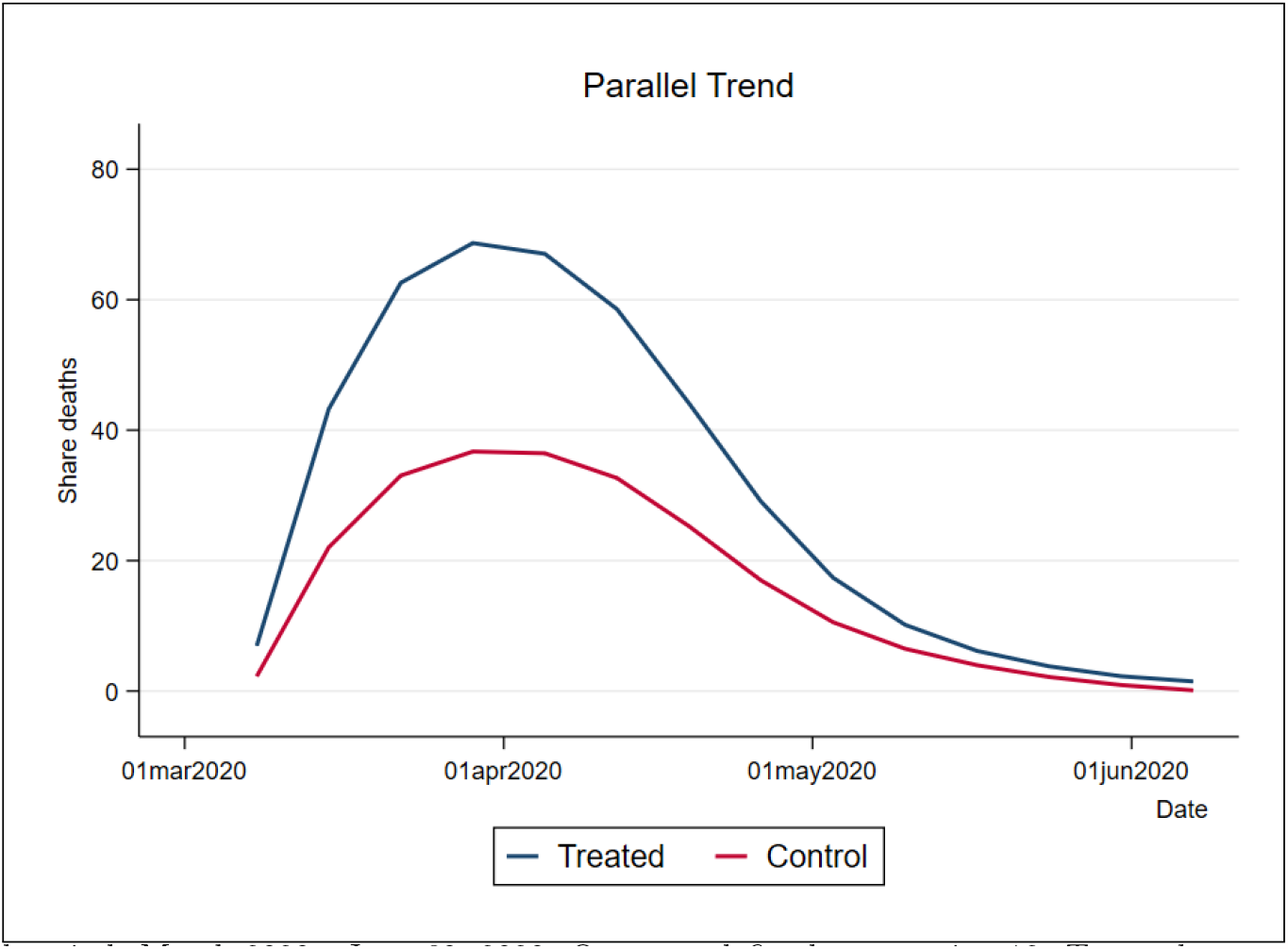
Parallel trends of Covid cases share Notes: Pre-trend period: March 2020 - June 30, 2020. Outcome defined as equation 12. *T reated* cantons (blue line) are those that between July, 7 and October, 4 have imposed any mask requirement other then Federal indications (e.g in supermarket, restaurants, open space): BS, FR, GE, JU, NE, SO, VS, VD, ZH. *Control* cantons (red line) contains the 17 left cantons.

**Figure B2:**
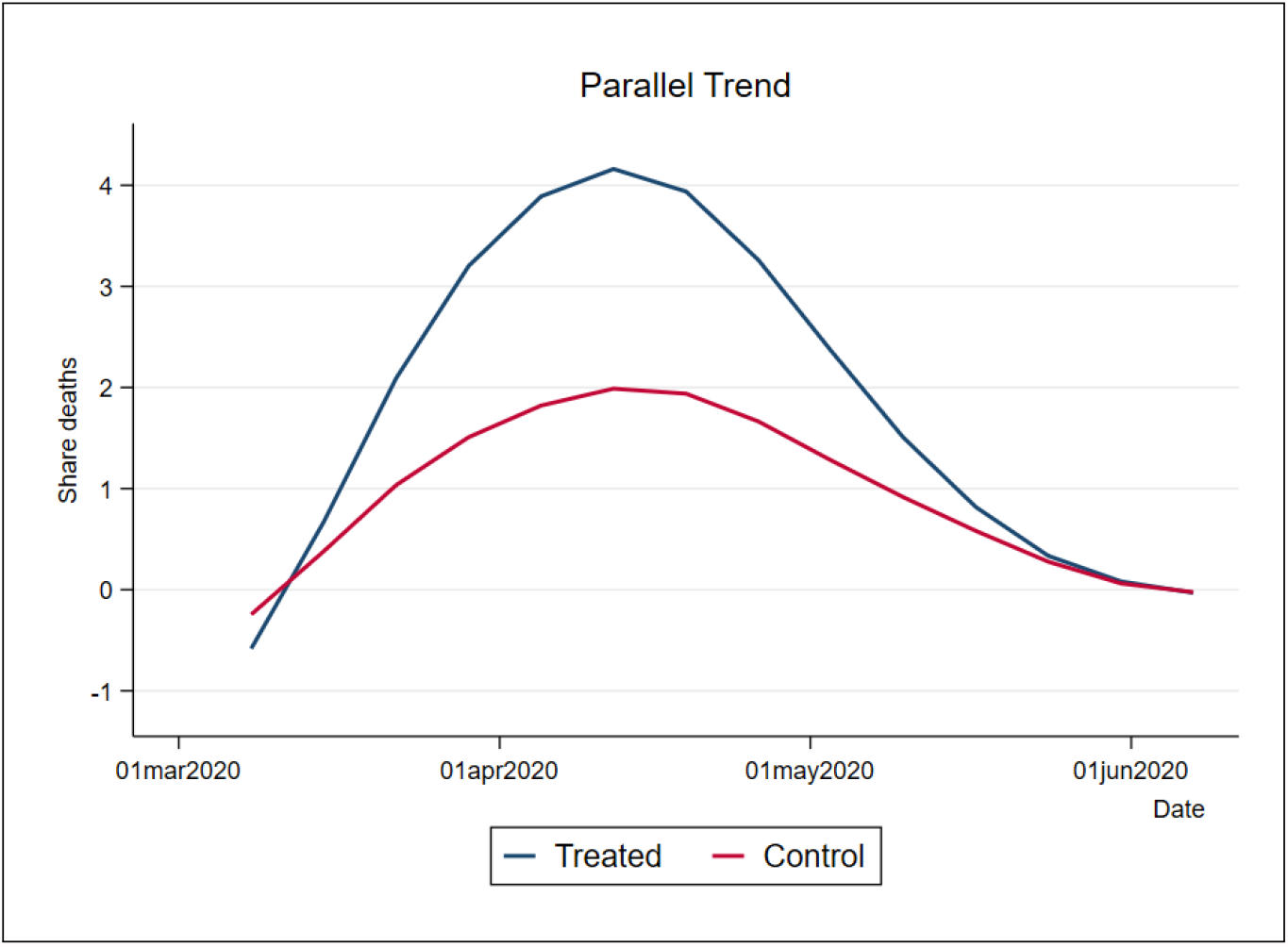
Parallel trends of Covid deaths Notes: Pre-trend period: March 2020 - June 30, 2020. Outcome defined as equation 13. *T reated* cantons (blue line) are those that between July, 7 and October, 4 have imposed any mask requirement other then Federal indications (e.g in supermarket, restaurants, open space): BS, FR, GE, JU, NE, SO, VS, VD, ZH. *Control* cantons (red line) contains the 17 left cantons.

**Table B2:**
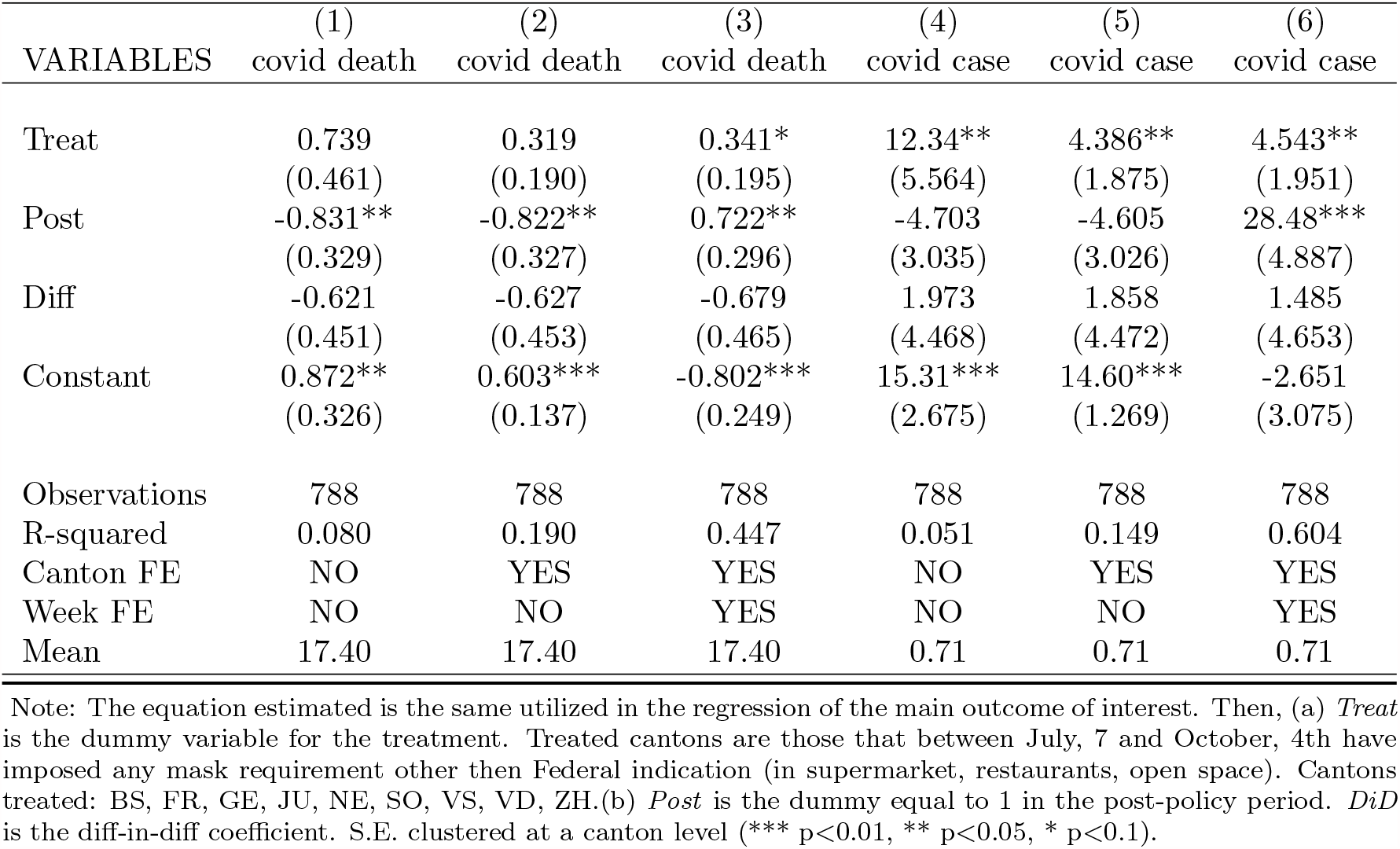
Difference-in-Difference regression on covid outcomes

| VARIABLES | (1)<br>covid death | (2)<br>covid death | (3)<br>covid death | (4)<br>covid case | (5)<br>covid case | (6)<br>covid case |
| --- | --- | --- | --- | --- | --- | --- |
| Treat | 0.739<br>(0.461) | 0.319<br>(0.190) | 0.341*<br>(0.195) | 12.34**<br>(5.564) | 4.386**<br>(1.875) | 4.543**<br>(1.951) |
| Post | -0.831**<br>(0.329) | -0.822**<br>(0.327) | 0.722**<br>(0.296) | -4.703<br>(3.035) | -4.605<br>(3.026) | 28.48***<br>(4.887) |
| Diff | -0.621<br>(0.451) | -0.627<br>(0.453) | -0.679<br>(0.465) | 1.973<br>(4.468) | 1.858<br>(4.472) | 1.485<br>(4.653) |
| Constant | 0.872**<br>(0.326) | 0.603***<br>(0.137) | -0.802***<br>(0.249) | 15.31***<br>(2.675) | 14.60***<br>(1.269) | -2.651<br>(3.075) |
| Observations | 788 | 788 | 788 | 788 | 788 | 788 |
| R-squared | 0.080 | 0.190 | 0.447 | 0.051 | 0.149 | 0.604 |
| Canton FE | NO | YES | YES | NO | YES | YES |
| Week FE | NO | NO | YES | NO | NO | YES |
| Mean | 17.40 | 17.40 | 17.40 | 0.71 | 0.71 | 0.71 |

Note: The equation estimated is the same utilized in the regression of the main outcome of interest. Then, (a) *Treat* is the dummy variable for the treatment. Treated cantons are those that between July, 7 and October, 4th have imposed any mask requirement other then Federal indication (in supermarket, restaurants, open space). Cantons treated: BS, FR, GE, JU, NE, SO, VS, VD, ZH. (b) *Post* is the dummy equal to 1 in the post-policy period. *DiD* is the diff-in-diff coefficient. S.E. clustered at a canton level (\*\*\* p<0.01, \*\* p<0.05, \* p<0.1).

**Figure B3:**
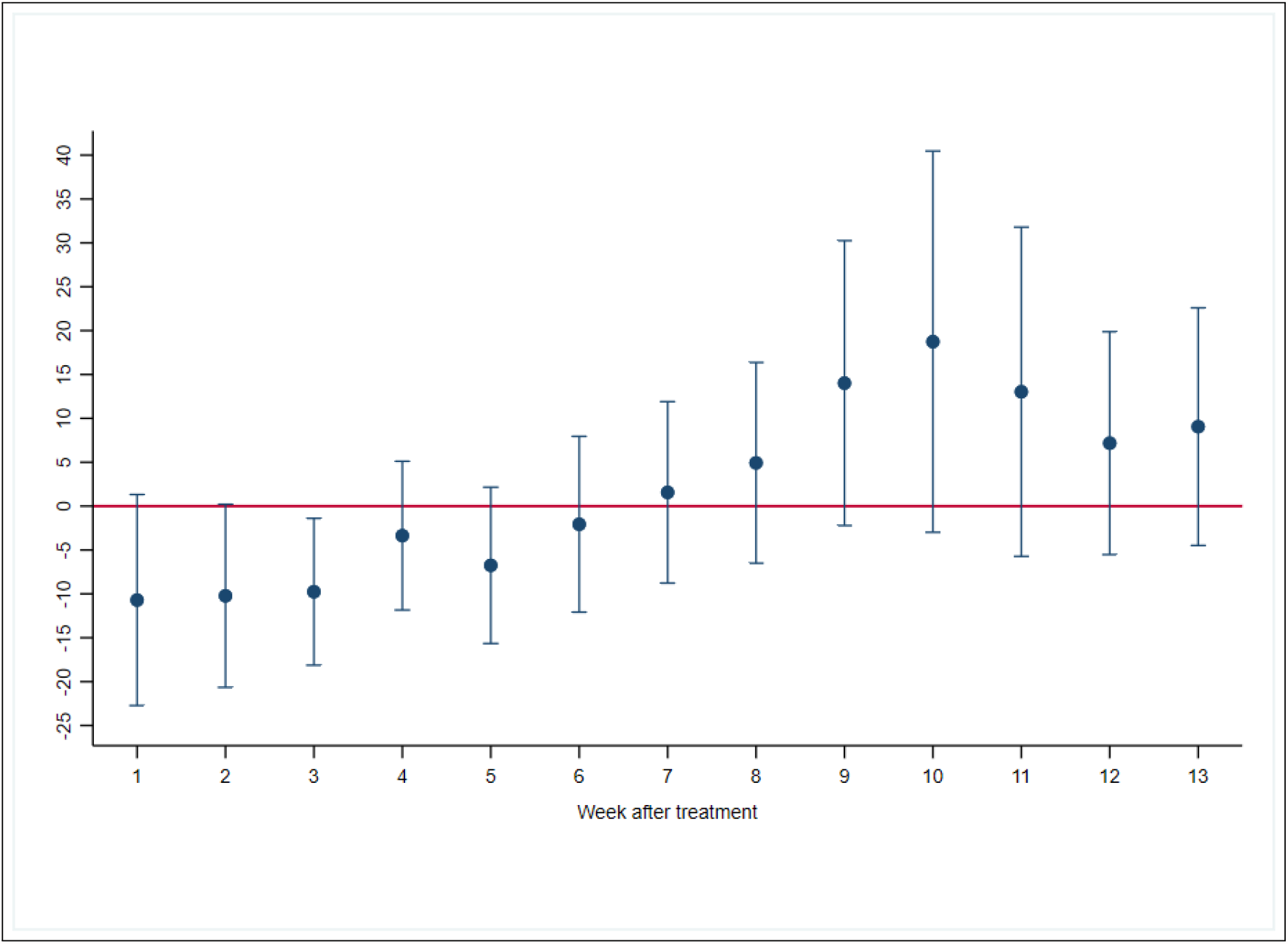
Difference-in-Difference regression on Covid cases share with dynamic Beta Note: Blue dots are the estimated *β*_2_ in week *w*. Week 1 is the first week after the treatment, until the 13^*t*^*h* week. Outcome defined as 12. Each bar represents the clustered SE of the point estimates (*** p*<*0.01, ** p*<*0.05, * p*<*0.1).

**Figure B4:**
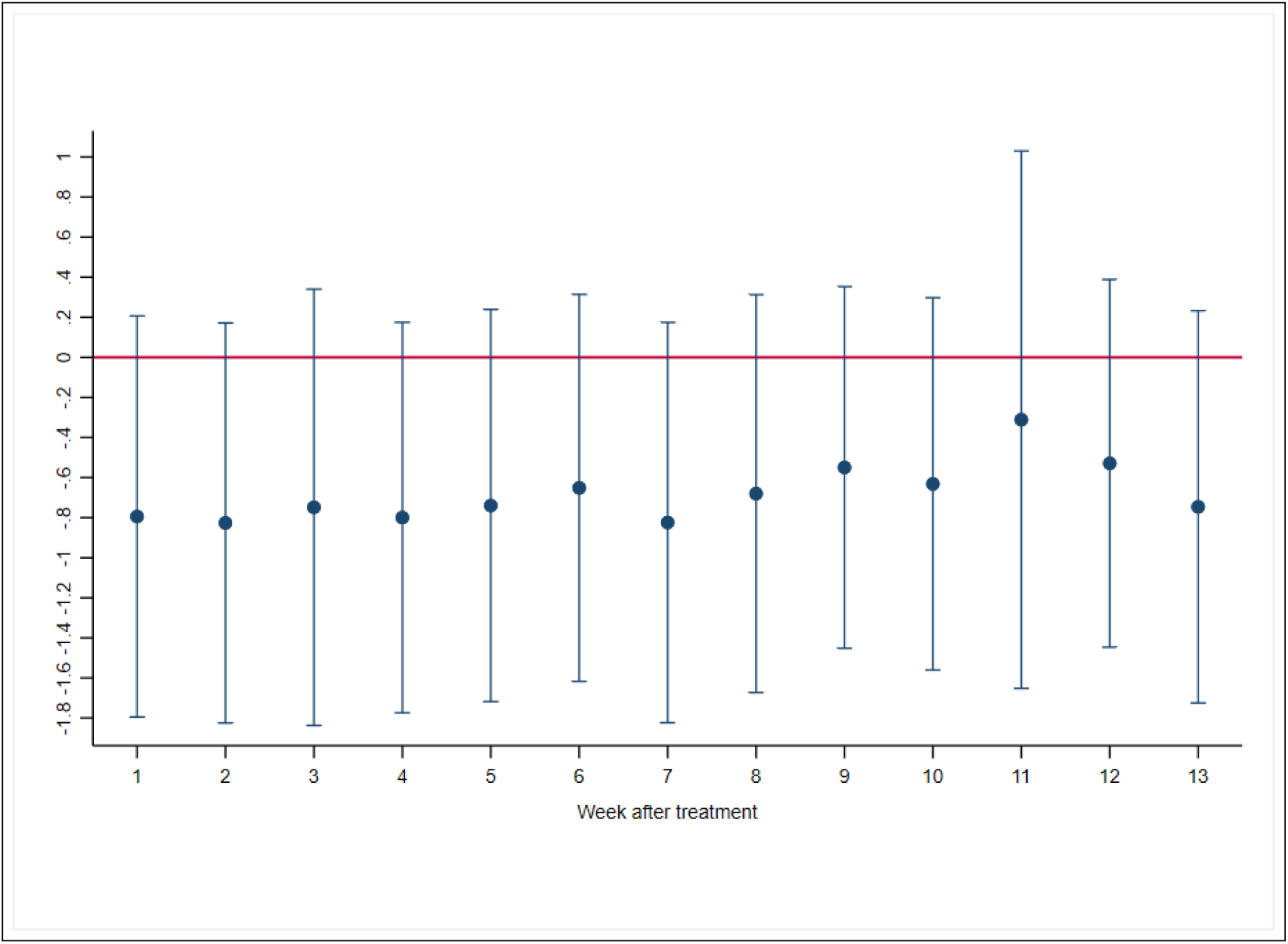
Difference-in-Difference regression on Covid deaths share with dynamic Beta Note: Blue dots are the estimated *β*_2_ in week *w*. Week 1 is the first week after the treatment, until the 13^*t*^*h* week. Outcome defined as 13. Each bar represents the clustered SE of the point estimates (*** p*<*0.01, ** p*<*0.05, * p*<*0.1).

## Footnotes

1 During this period, people were allowed to leave their houses and meet in groups of maximum 5 people, while all bars, clubs, shops and restaurants had to close. Only essential shops (e.g. food and beverages) and health facilities remained opened. In addition, borders with France, Austria and Germany were partly closed. These were re-opened between June 15 and June 22.

2 Starting from August 15 wearing the masks became compulsory on airplanes. https://www.bag.admin.ch/bag/en/home/krankheiten/ausbrueche-epidemien-pandemien/aktuelle-ausbrueche-epidemien/novel-cov/massnahmen-des-bundes.html

3 https://www.bfs.admin.ch/bfs/en/home.html

4 We keep in the main text the Tables where we verify parallel pre-trends by age-gender cohorts, but we also show the estimates of equation (3) on the whole period for 0-29, 60-90 and 90+ classes (tables (A3)-(A5)).

5 In Appendix A, tables (A11)-(A14) report the results of equation (10) estimation on the four age cohorts.

